# Stable Network-Level Functional Connectivity Alterations in Alzheimer’s Disease Identified via Interpretable Latent Modelling

**DOI:** 10.64898/2026.01.07.26343590

**Authors:** Santiago V. Blas Laguzza, Hugo Aimar, Diego M. Mateos, Martin A. Belzunce

## Abstract

Resting-state fMRI provides a non-invasive window into large-scale network-level alterations in Alzheimer’s disease (AD), but the high-dimensional functional connectivity (FC) and multi-site heterogeneity pose challenges to both classification and interpretability. We propose an explainable deep-learning framework that combines diagnosis-agnostic latent representation learning with a rigorously nested and interpretable classification pipeline to identify reproducible connectivity biomarkers of AD. Using a multi-site ADNI cohort (rs-fMRI, *N* = 431; 95 AD, 89 CN, 247 MCI), a diagnosis-agnostic *β*-VAE was trained diagnosis-agnostically on CN+MCI+AD data to learn a smooth latent representation of multichannel connectivity. We evaluated seven candidate connectivity measures spanning static, graph-filtered, dynamic, and effective connectivity. A systematic ablation study identified three complementary static channels as the most informative: Full Pearson correlation, OMST-reweighted Pearson, and *k*NN-based Mu-tual Information. Fold-specific latent means were then combined with age and sex and classified under a strictly nested 5 × 5 cross-validation scheme. Across outer test folds, Logistic Regression achieved a mean ROC–AUC of 0.843 and PR–AUC of 0.864. Pooled out-of-fold predictions for CN vs. AD subjects yielded a ROC–AUC 0.829 and a low Brier score (0.197), indicating well-calibrated probabilities. An attribution pipeline combining SHAP in latent space, followed by Integrated Gradients backprojected to edges, recovered a compact and reproducible AD signature. At the systems level, the differential saliency highlights Default Mode, Limbic, and Visual networks. Enforcing strict croos-fold stability criteria (replication frequency and sign consistency) yielded a consensus subgraph of 11 highly reliable connections. Importantly, the identified multivariate signature was largely disjoint from the strongest mass-univariate effects, indicating that the model captures complementary information beyond marginal group differences. Confounder analyses showed only moderate persistence of scanner-related signal in the latent space, with no evidence that acquisition or demographic factors systematically drove AD classification performance. Together, these results demonstrate a principled route toward interpretable, reproducible, and well-calibrated connectivity-based biomarkers for Alzheimer’s disease. All code, trained models, and mappings are publicly available to support transparency and external validation.

## 1 Introduction

Alzheimer’s disease (AD) is the most prevalent neurodegenerative disorder worldwide and represents a major public health challenge. It is characterized by progressive cognitive decline, memory loss, and behavioral changes that severely impair daily functioning and quality of life. Neuropathologically, AD is associated with the accumulation of amyloid-*β* plaques and tau protein tangles in the brain, leading to synaptic dysfunction and neuronal death Bloom [2014]. AD accounts for approximately 60–70% of all dementia cases and currently affects more than 55 million people globally, a number expected to rise dramatically as populations age Kamatham et al. [2024].

Beyond its molecular hallmarks, AD is increasingly understood as a disorder of large-scale brain networks. Structural and functional neuroimaging studies have consistently shown widespread alterations in inter-regional communication, particularly affecting long-range associations in the cerebral cortex. Resting-state functional MRI (rs-fMRI) provides a non-invasive means to probe these network-level abnormalities by measuring spontaneous correlations in blood-oxygen-level–dependent (BOLD) signals between brain regions. Numerous studies have reported AD-related disruptions in functional connectivity (FC), especially within the Default Mode Network (DMN), limbic circuits, and other association networks Wu et al. [2011]; Ibrahim et al. [2020]; Sanz-Arigita et al. [2010]; Supekar et al. [2008]. These alterations are detectable even at early disease stages and have motivated the use of rs-fMRI as a potential biomarker for AD diagnosis and progression monitoring.

However, translating rs-fMRI connectivity into reliable diagnostic tools remains challenging. Functional connectomes are high-dimensional, noisy, and sensitive to acquisition and preprocessing differences, particularly in large multi-site cohorts. Classical approaches based on mass-univariate statistics or handcrafted graph-theoretical metrics often suffer from limited sensitivity and poor generalization. Supervised machine-learning models can improve discrimination between cognitively normal (CN) individuals and AD patients, but they are prone to overfitting and instability, especially when trained directly on full connectomes Cawley and Talbot [2010]; Varoquaux [2018]; Dadi et al. [2019]. More recently, deep learning models have achieved promising classification performance, yet end-to-end architectures often entangle disease-related signal with site, scanner, or demographic confounds and provide limited interpretability, hindering biological insight and clinical trust Dinsdale et al. [2021]; Abrol et al. [2021].

Two key gaps therefore remain. First, there is a need for representations of functional connectivity that are both expressive and robust to non-disease variability, enabling generalisable classification across heterogeneous datasets. Second, beyond predictive accuracy, models must yield stable and biologically meaningful explanations, identifying reproducible connectivity patterns rather than fold-or cohort-specific effects. Addressing these challenges requires separating representation learning from supervised discrimination and enforcing stability at multiple stages of the analysis.

In this work, we propose a multistage framework that combines diagnosis-agnostic representation learning, strictly nested supervised classification, and fold-stable interpretability. We first learn a compact, non-linear embedding of multichannel functional connectivity using a convolutional *β*-Variational Autoencoder (*β*-VAE) trained without diagnostic labels. This latent representation is then used for CN–AD classification under a fully nested cross-validation protocol. To interpret model predictions, we integrate SHAP values computed in latent space with Integrated Gradients to map disease-relevant signal back to individual functional connections, and we explicitly quantify fold-wise stability to isolate reproducible effects. This design enables robust discrimination while yielding interpretable and stable network-level signatures of disease-related functional disruption.

## 2 Materials and Methods

In this study, we implemented a two-stage machine learning framework for AD classification and biomarker discovery (Fig. 1). In the first stage, subject-level representations of resting-state functional connectivity were learned using a convolutional *β*-VAE trained on multichannel connectivity matrices. In the second stage, supervised classifiers were applied to the resulting latent embeddings to discriminate CN individuals from patients with AD and to evaluate diagnostic performance.

**Figure 1:**
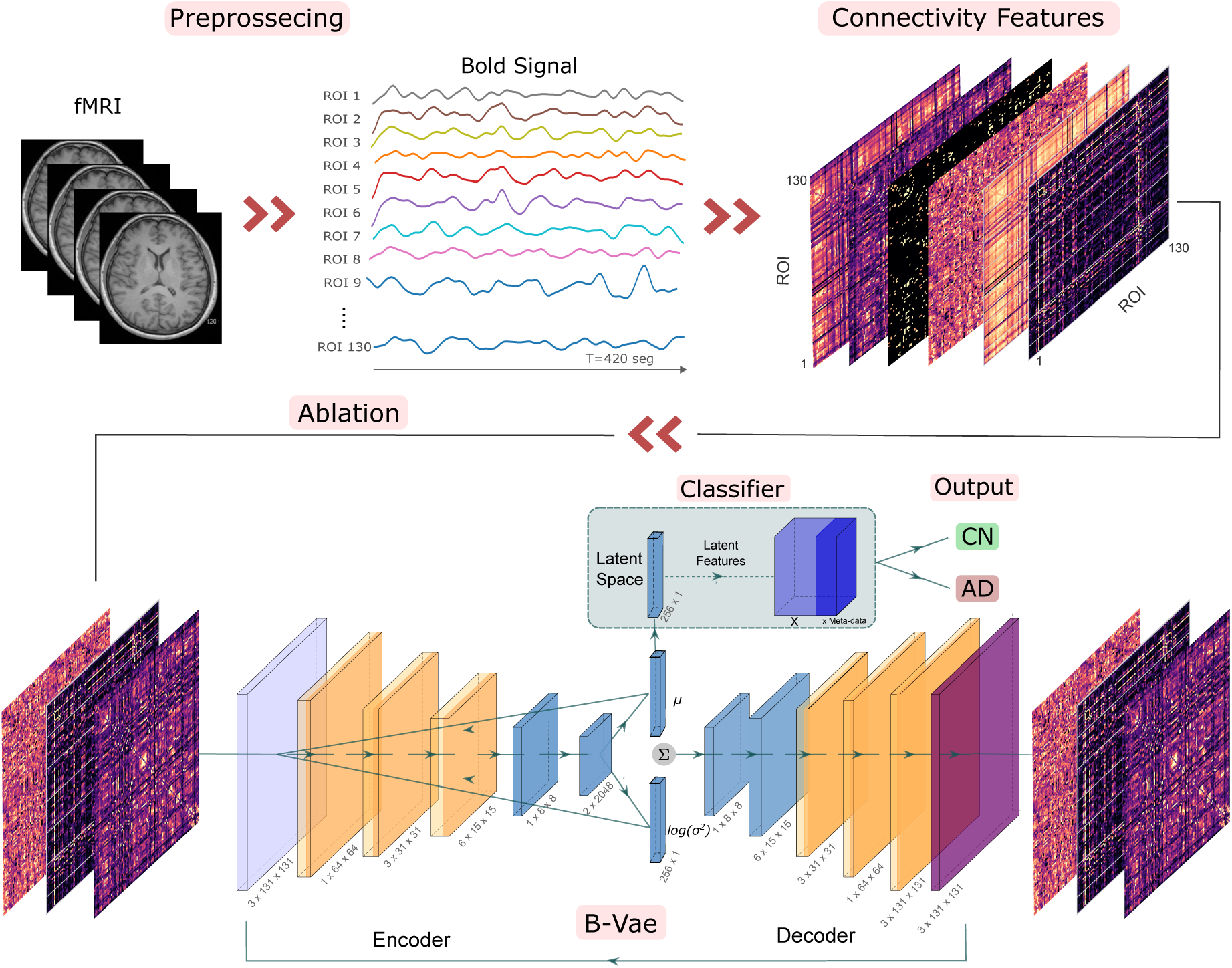
Two-stage pipeline. Resting-state functional connectivity matrices are first encoded into compact subject-level representations using a diagnosis-agnostic convolutional β-VAE trained within each outer cross-validation fold (including CN, MCI, and AD subjects). In the second stage, supervised classifiers are trained on the resulting latent embeddings, concatenated with age and sex, to discriminate CN from AD subjects under a fully nested 5×5 cross-validation scheme. Model interpretability is achieved by combining SHAP values in latent space with Integrated Gradients to project disease-related information back to individual connectivity edges.

All analyses were conducted under a fully nested cross-validation scheme to ensure leakage-free evaluation: an inner loop was used for model and hyperparameter selection, and an outer loop for unbiased performance assessment. Fold-specific encoders and classifiers were then integrated into an explainable AI (XAI) pipeline combining SHAP and Integrated Gradients, enabling the identification of stable and biologically interpretable connectivity biomarkers. Specifically, we used a 5×5 nested cross-validation design (5 outer folds, each with 5 inner folds).

### 2.1 Participants

We used resting-state functional MRI (rs-fMRI) data from the Alzheimer’s Disease Neuroimaging Initiative (ADNI)(adni.loni.usc.edu), specifically from the ADNI2, ADNI GO and ADNI3 cohorts. All images were acquired following the basic ADNI functional imaging protocol with a repetition time (TR) of 3 s. For each participant, we selected the rs-fMRI scan corresponding to the initial (baseline) visit.

The dataset included participants classified into three diagnostic groups: cognitively normal (CN), mild cognitive impairment (MCI, including both early [EMCI] and late [LMCI] subtypes), and Alzheimer’s disease (AD). After quality control, the final sample comprised *N* = 431 subjects. Regarding the study phase, 143, 11, and 277 subjects belonged to the ADNI2, ADNI GO, and ADNI3 cohorts, respectively. The demographic and key clinical characteristics of the study sample are summarised in Table 1. The CN/AD subset used for supervised classification comprised N=184 subjects (AD=95, CN=89).

**Table 1:** Demographic and clinical characteristics of the study cohort after quality control.

|  | AD | CN | MCI | Total |
| --- | --- | --- | --- | --- |
| Number | 95 | 89 | 247 | 431 |
| Age (years, mean ± SD) | 74.3 ± 7.5 | 74.4 ± 7.4 | 73.3 ± 8.0 | 73.8 ± 7.7 |
| Sex (Male/Female) | 55/40 | 37/52 | 133/114 | 225/206 |

All data were acquired following protocols approved by the institutional review boards of the participating ADNI sites, and written informed consent was obtained from all participants or their authorised representatives.

### 2.2 Data Acquisition and Pre-processing

As established by the ADNI basic acquisition protocol, rs-fMRI images were acquired using 3T scanners with a gradient echo planar imaging (EPI) sequence. The acquisition parameters were TR = 3 s, TE = 30 ms, flip angle = 90°, matrix size 64 × 64 × 48, and voxel size 3.4 × 3.4 × 3.4 mm^3^.

The duration of the scans was 7 min for ADNI2 and 10 min for ADNI3, which resulted in 140 and 200 time points, respectively. Of the 431 images, 274 were acquired using Philips, 110 using Siemens, and 47 using GE scanners.

The rs-fMRI data were preprocessed with the MATLAB (The MathWorks, Inc., Natick, MA) toolbox “Data Processing Assistant for Resting-State fMRI” (DPARSF) Yan [2010], built upon SPM12 (Statistical Parametric Mapping, Wellcome Trust Centre for Neuroimaging, London, UK, https://www.fil.ion.ucl.ac.uk/spm/software/spm12/), supplemented with MATLAB in-house scripts to do the image quality check and to extract the BOLD signals for different atlases.

The pre-processing pipeline included slice timing correction (adjusted for each scanner configuration), realignment for motion correction, spatial normalisation to the Montreal Neurological Institute (MNI) standard space, spatial smoothing with an isotropic Gaussian kernel with a full-width at half-maximum (FWHM) of 4 mm, temporal bandwidth filtering with cut-off frequencies of 0.01-0.10 Hz, and nuisance covariate regression. All images were resampled to an isotropic voxel resolution of 2 × 2 × 2 mm^3^ during spatial normalisation. For the nuisance covariate regression, we regressed six motion parameters derived from the realignment step and mean signals extracted from the subject-specific white matter (WM) and cerebrospinal fluid (CSF) masks.

After DPARSF processing, we applied grand mean intensity normalisation to rescale the global brain signal to a mean value of 10, 000. To ensure data quality, we computed the signal-to-noise ratio (SNR) and temporal signal-to-noise ratio (tSNR) for each scan. Based on these metrics and a comprehensive quality control report, 3 subjects were excluded due to preprocessing errors or poor image quality (we had 434 patients originally).

Finally, regional time series were extracted by parcellating the preprocessed fMRI data using the Automated Anatomical Labelling atlas version 3 (AAL v3) Rolls et al. [2020]. We initially obtained the mean BOLD signal for the 170 ROIs of the AAL3 atlas and then excluded four empty regions and 35 additional ROIs with fewer than 100 voxels to avoid instability, resulting in a final set of 131 ROIs. To ensure a uniform input length for all subsequent analyses, each time series was homogenized to a length of 140 time points by truncating longer series. Each ROI time series was *z* -scored (zero mean, unit variance).

The 131 resulting ROIs were then reordered according to the Yeo-17 functional network template Thomas Yeo et al. [2011]. To achieve this, the AAL3 atlas was resampled to the same spatial reference as the Yeo-17 atlas, and each ROI was assigned to the Yeo network with the greatest spatial overlap. This reordering grouped functionally related regions (*e.g.,*visual, default-mode) into contiguous blocks in the connectivity matrices. Organizing the matrices in this way facilitates the detection of within-and between-network connectivity patterns and the interpretation of model outputs in terms of established functional brain systems. The complete ROI-to-network mapping is provided in the public repository listed under the Data and Code Availability section.

Finally, for each participant, we obtained a matrix **Y** ∈ ℝ^140×131^ whose columns represent the harmonized BOLD signals from the 131 ROIs, arranged in Yeo-aligned functional order. This matrix was then used for the multichannel connectivity analysis described next.

### 2.3 Functional Connectivity Feature Extraction

Analyzing functional connectivity in fMRI data is essential for characterizing brain network alterations in Alzheimer’s disease. Because no single metric can fully capture the complexity of inter-regional dependencies, we initially evaluated a comprehensive set of seven candidate channels:

1. *Full Pearson (Fisher z).* Dense Pearson correlation between ROI series, Fisher *z*-transformed (sign preserved).
2. *Orthogonal Minimum Spanning Tree (OMST)-Pearson.* Sparse backbone (binary OMST mask on |*r*|) re-weighted with signed Fisher *z*.
3. *Distance Correlation.* It captures linear and nonlinear dependencies via pairwise distance structures.
4. *Mutual Information kNN.* Symmetric MI estimated with *k*=5 nearest neighbours.
5. *Dynamic FC: mean absolute fluctuation.* Mean |ΔFC| from sliding windows (30 TR, step 5).
6. *Dynamic FC: temporal instability.* Across-window variability of FC (same scheme).
7. *Symmetric Granger F-scores.* A proxy for effective connectivity (lag = 1).

Each of the seven candidate channels is represented by a 131×131 matrix, with the diagonal zeroed and off-diagonal entries *z*-scored (fitted only to the training set). Metrics 1–4 were computed for the full time series. Metrics 5 and 6 used a sliding-window approach (90 s windows, 15 s steps), where we computed the Pearson correlation for each window and then applied the dynamic connectivity metrics to the resulting set of correlation matrices. For details on connectivity metrics, see the Supplementary Section S1 "Connectivity measures: definitions and implementation details".

An ablation study was performed to select the number of channels (*N_C_*) and determine which of them achieved higher classification performance. This process was performed first with an initial lightweight analysis, where we evaluated the individual and combined predictive value of all candidate channels using a greedy forward selection. This analysis was later re-evaluated with the final high-capacity VAE framework.

### 2.4 VAE-based Representation Learning

To learn a compact, non-linear embedding of the high-dimensional connectomes, we trained a convolutional *β*-Variational Autoencoder (*β*-VAE) Higgins et al. [2017]. The encoder mapped each subject’s input tensor C ∈ ℝ*^NC^* ^×131×131^ to a 256-dimensional latent distribution. The mean vector *µ* ∈ ℝ^256^ of this distribution served as the subject-level embedding for the subsequent classification. The *β*-VAE was optimized using the standard Evidence Lower Bound (ELBO), where the reconstruction loss is regularized by a Kullback–Leibler divergence term weighted by *β*. The training parameter selection included a cyclical annealing schedule for the *β* hyperparameter with 32 cycles (final *β* = 2.5) and the AdamW optimizer with a cosine annealing with warm restarts learning rate scheduler (T_0_=120 epochs, *η*_min_ = 5 × 10^−7^).

The network consisted of four convolutional blocks with GELU activations, group normalization, and dropout (rate 0.2), mirrored by a symmetric decoder using transposed convolutions.

The VAE was trained for a maximum of 2400 epochs using a batch size of 64, with early stopping patience set to 120 epochs on the validation loss. Within each outer cross-validation fold, the VAE was trained diagnosis-agnostically only on the corresponding training pool (including CN, MCI, and AD) and never saw the held-out test set, ensuring a leakage-free representation learning. The full architecture and training details are provided in the Supplementary Section S2 "Architecture and training".

To characterise how the VAE structured the connectomic manifold, we computed several quantitative metrics on the held-out test subjects of each outer fold:

- Class separability: Silhouette score computed on a 20-dimensional PCA projection of the latent space.
- Intrinsic dimensionality: Participation Ratio (PR) computed from the covariance spectrum of the latent means.
- Confounder decodability: Balanced accuracy of predicting scanner manufacturer or acquisition site using a shallow *k* -NN classifier trained only on training-fold latents.
- Demographic leakage: Generalisation performance of linear models trained to predict age or sex from training-fold latents and evaluated on the corresponding test fold.

After training, the encoder was frozen and the latent mean vectors *µ*(*x*) were extracted for all subjects. These latent embeddings were the only imaging features used by the supervised classifiers of the next section.

### 2.5 Supervised Classification

Supervised classification was performed exclusively on the CN vs. AD subset (*N* = 184; 95 AD / 89 CN). For each participant, the 256-dimensional latent mean vector *µ* from the corresponding fold-specific encoder was concatenated with {Age, Sex} covariates to form the input feature set.

We then trained supervised classifiers to discriminate between cognitively normal (CN) and Alzheimer’s disease (AD) subjects.

We evaluated five algorithmic families covering linear, kernel-based, and ensemble methods: a small MLP, regularised Logistic Regression, RBF-SVM, XGBoost, and Gradient Boosting (GB). All steps were wrapped in a single imblearn pipeline. We applied StandardScaler to the non-tree models. To mitigate class imbalance, we used class weighting within the inner folds. We did not apply synthetic oversampling (no SMOTE) in the final reported experiments.

Model selection used a fully nested cross-validation scheme: 5-fold outer CV for unbiased evaluation and a 5-fold inner CV for hyperparameter optimisation with Optuna (TPE sampler, MedianPruner), maximizing ROC–AUC. Performance on each outer test fold was quantified by ROC–AUC (primary metric), PR–AUC, balanced accuracy, sensitivity, specificity, and F_1_, summarised as mean ± SD across outer folds. The VAE encoder was always trained excluding the outer test subjects, preventing leakage and guaranteeing strict separation between representation learning and classification.

Additional details, including search spaces, runtime parameters, and configuration scripts, are provided in the public repository listed in the Data and Code Availability section and in the Supplementary Sections S3 "Supervised classification pipeline" and S4 "End-to-end workflow (summary)".

### 2.6 Interpretability Framework for Biomarker Discovery

To move beyond classification and identify biologically interpretable connectivity biomarkers, we implemented a two-stage explainable AI framework that links model predictions to the input connectomes. In the first stage, the SHAP values are used to determine which latent dimensions of the VAE space are most relevant for AD classification. In the second stage, integrated gradients are applied to backpropagate the AD-related latent dimensions to every edge of the connectome Sundararajan et al. [2017]. This yields a saliency map indicating which connections consistently contribute to the AD-related signal.

#### Controlling demographics in SHAP

All interpretability analyses use the *same* classifier trained in the main pipeline (input = [***µ*** +Age+Sex]; no retraining). We distinguish two SHAP interrogation modes applied *after* training: *unfrozen*, where Age/Sex vary naturally; and *frozen*, where Age and Sex are replaced by their training-set statistics (mean/mode) during SHAP sampling, removing their variance and forcing their SHAP contribution to zero. Formally, letting *z* = [*µ*_1_*, . . . , µ*_256_, Age, Sex], the frozen mode evaluates SHAP on *z̃* with Age= *a*ā_train_ and Sex= *s̄*_train_, isolating connectivity-driven attributions. Unless stated otherwise, all biomarker results (saliency maps, consensus edges, hubs) are derived from the *frozen* setting. The *unfrozen* setting is reported as a sensitivity check in the Supplement (where Age emerges as the top feature), and it yields virtually identical rankings within the 256 latent dimensions (mean Pearson *r >* 0.999 across folds).

#### Stage 1: SHAP in latent space

For each trained classifier of the outer folds, we computed SHAP values (SHapley Additive exPlanations) *φ_ℓ_*(*x*) for each *ℓ*-th latent feature (*µ* ∈ ℝ^256^ plus age and sex) of a subject *x* Lundberg and Lee [2017]. To quantify the overall importance of each latent feature, we computed the group-contrast weights *w_ℓ_* by calculating the difference in the mean SHAP values between the two groups, as follows:

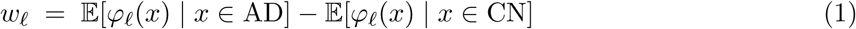

To focus only on relevant brain signatures, we define a refined *L*_1_-normalised weight vector *w̃*, where we keep the Top-*K* latent coordinates (*K*=50) ranked by |*w_ℓ_*| and set the remaining weights and demographic covariates (age and sex) to zero:

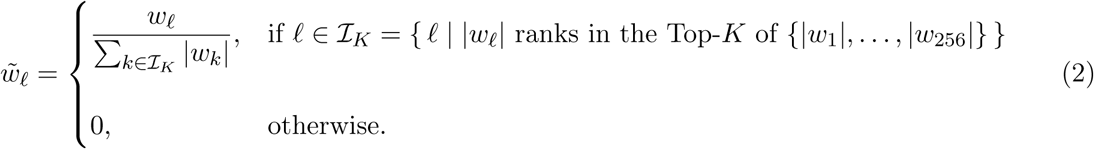

This yields an AD-oriented score in the latent space for each subject *x*, defined as a linear function of the mean vector *µ*(*x*):

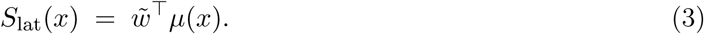

#### Stage 2: Integrated Gradients back-projection

Using the frozen encoder and SHAP-derived weights, we computed Integrated Gradients (IG) to attribute the AD-oriented score to the input connectivity features across the *N_C_* channels. The baseline connectome was defined as the channel-wise median of CN subjects of matching age and sex from the training pool of each fold, providing an on-manifold reference representing “absence of disease signal.”

To propagate the AD-oriented latent score back to the input connectome, we compose the latent scoring function with the encoder to define an input-dependent scalar function as follows:

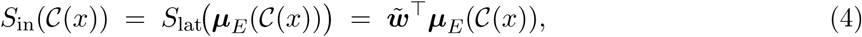

where *µ_E_*(·) denotes the encoder latent mean mapping. Although *S*_in_ is computed through the latent representation, it remains a differentiable scalar function of the input connectome C(*x*), allowing gradient-based attribution with respect to each edge (*i, j*) via IG. Note that *S*_in_ is not the classifier logit or probability, but a SHAP-weighted latent score that we deliberately construct to bridge latent and input spaces in a controlled way.

For each subject *x*, we denote by *S_ij_*(*x*) the saliency assigned to edge (*i, j*) by the Integrated Gradients back-projection (after symmetrising and aggregating across channels). To summarise group differences at the edge level, we define the AD–CN *differential saliency* as

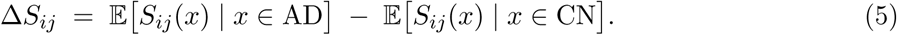

Δ*S_ij_ >* 0 indicates a *pro-AD* edge (its deviations from the CN baseline increase the AD-oriented score *S*_in_), whereas Δ*S_ij_ <* 0 indicates a *pro-CN* edge. Thus, Δ*S_ij_* quantifies how much more (or less) an edge contributes to *S*_in_ in AD than in CN, rather than a direct projection of latent vectors onto a disease direction. All edges are ranked by |Δ*S_ij_*|.

For completeness, we computed fold-wise summaries of channel contributions by aggregating absolute Integrated Gradients across edges and subjects for each connectivity channel.

#### Stability and consensus across folds

To ensure the robustness of the edge-level saliency maps obtained from Integrated Gradients, we assessed their stability across outer folds. For each outer fold *f* , edges were ranked by absolute differential saliency. Let 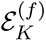 denote the Top-*K* edges in fold *f* . For each edge (*i, j*), we define its replication frequency and mean signed direction across F folds as

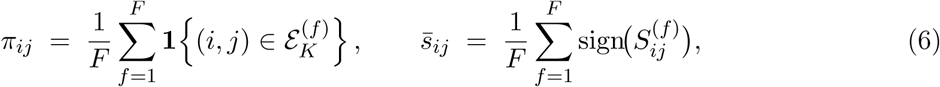

where 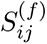 denotes the fold-specific differential saliency for edge (*i, j*).

A signed consensus weight combining stability and directionality was then defined as

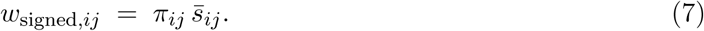

Edges were retained in the consensus set if *π_ij_* ≥ 0.6 and showed consistent direction across folds (|*s̄_ij_*| ≥ 0.6). Average pairwise Dice overlaps and Spearman correlations between folds were also computed as complementary stability metrics.

For interpretability, we summarized the edge-level consensus saliency at the functional-network level using the Yeo–17 template. Specifically, for each pair of networks, we aggregated the signed (and absolute) differential saliency of all consensus edges connecting regions within or between those networks, yielding a systems-level representation of the AD-related connectivity signature.

#### Hemispheric patterns

To test hemispheric asymmetry, we first filtered the saliency rankings to include only intra-hemispheric connections (excluding midline ROIs). For a given threshold *K* (e.g., *K*=50, 100, 200), we pooled the counts of right-right (R-R) and left-left (L-L) connections from the Top-*K* lists across all five folds. We then performed a one-sided binomial test on these aggregated counts against the null hypothesis of equal probability (*H*_0_ : *p*_RR_ = 0.5) to assess symmetry. As a robustness check, we repeated the test per fold and combined p-values via Fisher’s method, reaching the same conclusion.

#### Saliency–effect size linkage for validation

To assess whether model-derived saliencies correspond to genuine group differences, we compared edge-wise IG attributions with empirical effect sizes (Cohen’s *d*; (*d_ij_*) computed on held-out test data. We first derived a consensus set from saliency and, independently, a consensus set from Cohen’s *d* (using the same fold structure). As a sensitivity analysis, we examined the relation both within the 11-edge saliency consensus and on pooled Top-*K* (non-consensus) edges.

#### Reproducibility and leakage control

All SHAP and IG computations employed the fold-specific encoder 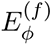 frozen at the validation-selected epoch. The baselines and scalers were fitted exclusively on the training data within each fold. All experiments were implemented in Python using PyTorch and scikit-learn on a single NVIDIA GPU (CUDA12.1). The full code, models, and results are publicly available to guarantee reproducibility (see the Data and Code Availability section).

## 3 Results

We organise the results in four steps. We first describe the dataset and nested cross-validation design. We then summarise the connectivity-channel ablations that motivated a three-channel configuration. Next, we characterize the structure and confounder content of the *β*-VAE latent space. Finally, we report CN–AD classification performance and the stable connectivity signature recovered by the SHAP→IG explainability pipeline.

### 3.1 Dataset and cross-validation overview

After preprocessing and quality control, the final dataset comprised *N* = 431 subjects from ADNI2, ADNI GO and ADNI3 (95 AD, 89 CN, 247 MCI). Within each outer fold, the *β*-VAE was trained diagnosis-agnostically on the full training pool (CN+MCI+AD), ensuring that representation learning did not use diagnostic labels.

Supervised classification and interpretability analyses were restricted to CN and AD subjects. This resulted in *N* = 184 participants (95 AD, 89 CN) entering the outer folds of the nested protocol, with each subject acting exactly once as test data. All performance metrics reported below are based on strictly out-of-fold predictions: neither the VAE nor the classifiers ever saw test subjects during training, model selection, or hyperparameter optimisation.

### 3.2 Connectivity channel ablation and selection

We first performed a lightweight ablation study to evaluate the predictive value of each candidate connectivity channel in isolation and in small combinations. In the single-channel analysis, Full Pearson (Fisher-*z*) emerged as the strongest individual predictor (mean ROC–AUC = 0.75 ± 0.02), whereas the remaining channels yielded substantially lower performance. A greedy forward selection over channel combinations showed that predictive performance peaked at the two-channel combination Full Pearson + OMST-Pearson (mean ROC–AUC = 0.79 ± 0.03 standard error) (see Fig. 2). A more detailed explanation of this process is given in Supplementary Section S5 "Connectivity channel ablation".

**Figure 2:**
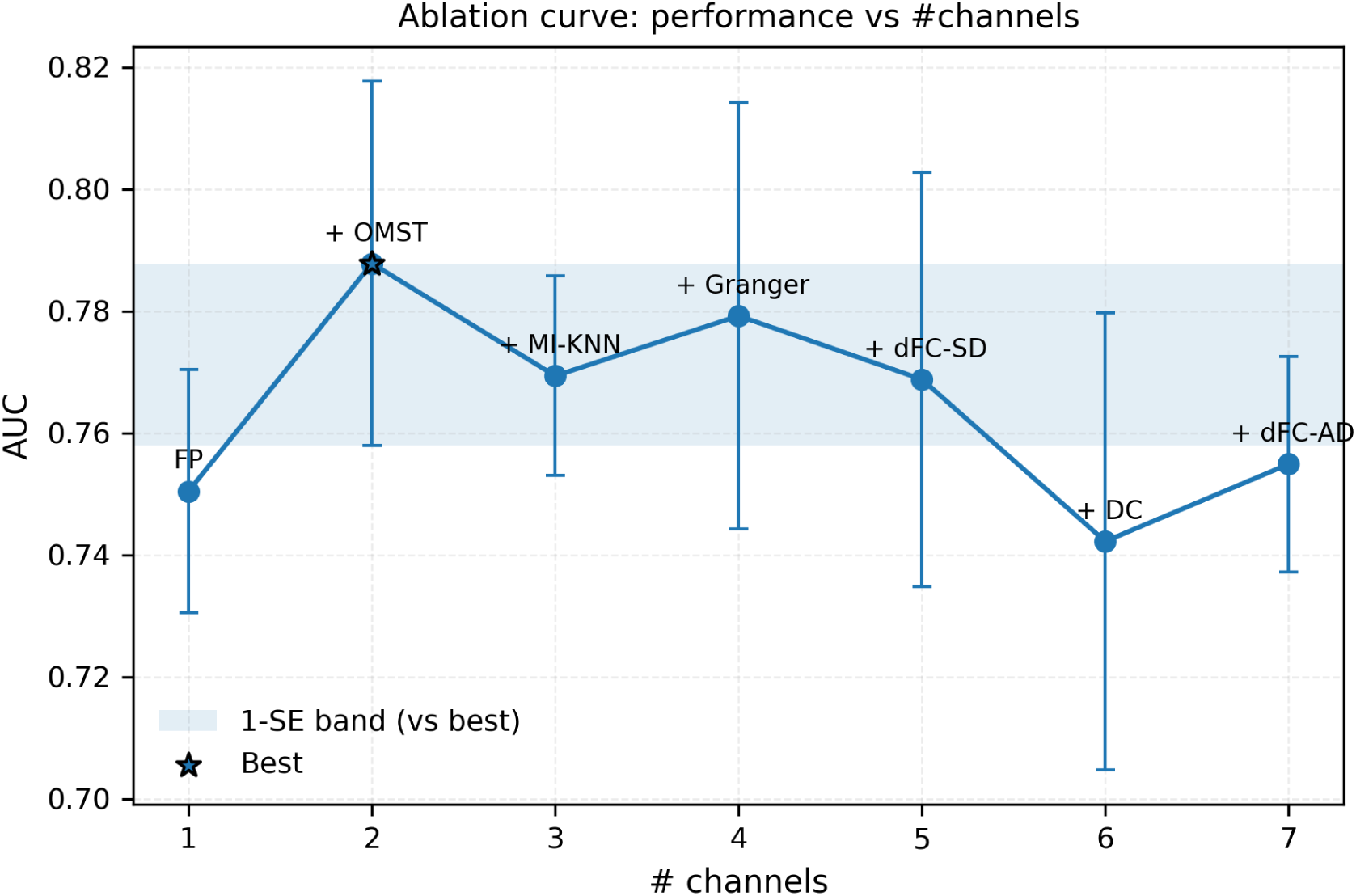
Greedy channel ablation curve (FAST configuration). Mean outer-fold ROC–AUC as a function of the number of connectivity channels included in the model. The candidate channels were: Full Pearson (Fisher-z) (FP), OMST–Pearson (Fisher-z weighted) (OMST), Mutual Information k-NN (MI-KNN), dynamic FC: mean absolute fluctuation (dFC-MAD), dynamic FC: temporal instability (dFC-SD), Distance Correlation (DC), and symmetric Granger F-scores (Granger). Error bars indicate the standard error across the three outer folds. The shaded region denotes the one-standard-error (1-SE) band relative to the best-performing model (*). Labels above points indicate the channel added at each step of the greedy forward-selection procedure.

When this analysis was re-evaluated in the full setting, using the high-capacity convolutional *β*-VAE and the complete nested classification pipeline, the three-channel tensor (Full Pearson, OMST-Pearson, Mutual Information *k*NN) consistently outperformed the two-channel model across outer folds. Based on this evidence, all subsequent VAE training, classification, and interpretability analyses were performed on three-channel connectivity tensors.

### 3.3 Latent representation learning with the **β**-VAE

Within each outer fold, a convolutional *β*-VAE was trained on three-channel connectivity tensors from the corresponding training pool (CN, MCI, and AD). The encoder produced 256-dimensional latent mean vectors, with a cyclical annealing schedule for the KL weight (final *β* = 2.5). Training converged stably in all folds, with smooth ELBO trajectories and no evidence of posterior collapse.

Although the latent dimensionality was set to 256, the effective intrinsic dimensionality, quantified via the Participation Ratio, was substantially lower and ranged from 18.7 to 21.0 across folds (Supplementary Table S2). Class separability in latent space was negligible: a Silhouette score computed on a 20-dimensional PCA projection yielded a mean of 0.005 ± 0.007, confirming that the VAE learned a general functional manifold without forming diagnosis-specific clusters.

We next quantified whether acquisition factors persisted in the latent representation. A *k*-NN confounder decoder trained on the standardised latent means was used to predict the three scanner manufacturers present in the dataset (chance level = 1*/*3 ≈ 0.33). Across outer folds, this decoder achieved a mean balanced accuracy of 0.42 ± 0.06 (range 0.33–0.50; Supplementary Table S2), indicating moderate manufacturer-related structure in the latent space. As a complementary analysis, a multinomial Logistic Regression decoder trained either on the *z*-scored input connectomes or on the latent means *µ* recovered the same scanner labels well above chance (balanced accuracy 0.678 ± 0.058 vs. 0.735 ± 0.054, respectively; Supplementary Table S2). Decodability remains of similar magnitude before and after VAE encoding, indicating that the VAE does not artificially suppress or exaggerate scanner-related structure.

We also quantified demographic leakage. Age-only regression models trained on the latent means explained none of the variance in held-out test data (test-set *R*^2^ consistently below zero)(i.e., worse than predicting the mean), and Sex-only classifiers reached an accuracy of 0.58 ± 0.14, only slightly above the 0.5 chance level. Importantly, none of the confound indices we analysed (latent-space class separability, manufacturer/site decodability, or fold-wise summaries of age and sex) showed a significant association with AD classification performance: Spearman correlations between outer-fold ROC–AUC and confound metrics ranged from −0.40 to 0.70, with all *p >* 0.10 (Supplementary Material S6 "Quantitative and Visual Validation of the Latent Space"). Together, these results indicate that the *β*-VAE produces a stable and structurally interpretable latent space that preserves inter-subject connectomic variability and disease-relevant axes, while keeping acquisition-related structure at a moderate level comparable to the input connectomes and with limited evidence that these confounds drive CN–AD discrimination.

### 3.4 AD classification performance

Classification between AD and CN was performed using the fold-specific 256-dimensional latent embeddings concatenated with age and sex, under a fully nested 5×5 cross-validation protocol. Among the evaluated classifiers, Logistic Regression achieved the highest outer fold performance (Table 2), with a mean ROC–AUC of 0.843 ± 0.069 and a mean PR–AUC of 0.864 ± 0.056.

**Table 2:** Nested 5-fold cross-validation performance (mean ± SD). The best performing model (Logistic Regression) is highlighted in bold.

| Model | ROC-AUC | PR-AUC | Bal. Acc. | Sens. | Spec. | F1 Score |
| --- | --- | --- | --- | --- | --- | --- |
| MLP | $0.78 \pm 0.12$ | $0.81 \pm 0.10$ | $0.68 \pm 0.10$ | $0.75 \pm 0.16$ | $0.62 \pm 0.34$ | $0.71 \pm 0.04$ |
| **LOGREG** | **$0.84 \pm 0.07$** | **$0.86 \pm 0.06$** | **$0.79 \pm 0.05$** | **$0.80 \pm 0.12$** | **$0.78 \pm 0.09$** | **$0.79 \pm 0.06$** |
| RBF-SVM | $0.81 \pm 0.06$ | $0.83 \pm 0.04$ | $0.73 \pm 0.04$ | $0.71 \pm 0.07$ | $0.75 \pm 0.05$ | $0.73 \pm 0.05$ |
| XGB | $0.81 \pm 0.06$ | $0.82 \pm 0.05$ | $0.72 \pm 0.05$ | $0.77 \pm 0.14$ | $0.66 \pm 0.10$ | $0.73 \pm 0.07$ |
| GB | $0.80 \pm 0.04$ | $0.81 \pm 0.04$ | $0.71 \pm 0.02$ | $0.74 \pm 0.05$ | $0.69 \pm 0.04$ | $0.73 \pm 0.02$ |

Pooling all out-of-fold predictions (*N* = 184; 95 AD / 89 CN), the model obtained a ROC–AUC of 0.83 and a Brier score of 0.197, indicating reasonably good probability calibration. The 95% bootstrap confidence intervals (2,000 resamples) for the pooled ROC and PR curves are shown in Fig. 3. The sharp transition in the PR curve reflects the strongly bimodal distribution of predicted probabilities across folds, consistent with good class separation rather than a thresholding artefact. Clinically relevant operating points, including Youden’s index, balanced-sensitivity threshold, high-sensitivity screening threshold, and a cost-sensitive rule, are reported in Supplementary Table S4.

**Figure 3:**
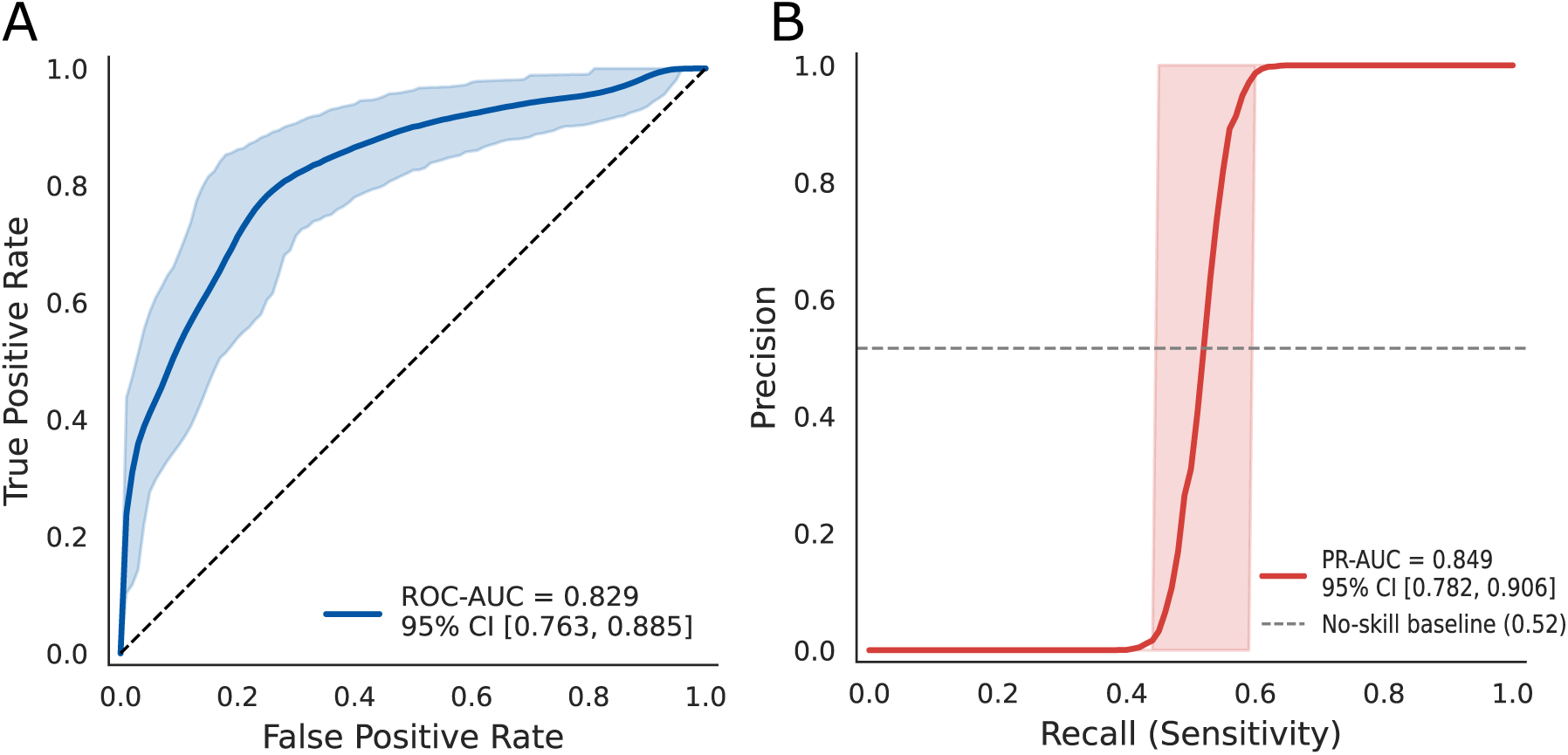
Classification performance of the cross-validated pipeline (LOGREG). **(A)** Pooled out-of-fold ROC curve (all test subjects concatenated). **(B)** Pooled out-of-fold Precision–Recall curve. Shaded areas: 95% bootstrap CIs (B=2000). Legends report pooled AUC and 95% CI; fold-wise mean AUCs are reported in Table 2.

These thresholds were computed *post hoc* on the pooled out-of-fold predictions and were not used for model selection.

Overall, these results demonstrate that the combined *β*-VAE representation and supervised classifier offer robust discrimination between AD and CN, based solely on multichannel functional connectivity.

### 3.5 Interpretability: a compact and stable multivariate connectivity signature

After computing SHAP scores in latent space (see Supplementary Material S8 "Global latent-space explanations (SHAP)") and projecting them back to the input connectomes using Integrated Gradients as described in methods section 2.6, we obtained fold-specific saliency maps that quantify the contribution of each functional connection to the AD–CN decision. Our interpretable deep learning pipeline reveals a stable and neurobiologically coherent functional connectivity signature of Alzheimer’s disease (AD), summarised in Fig. 4. In terms of channel contributions, they were fairly even, with contributions of 34%, 36% and 28% for Full Pearson, Mutual Information *k*NN and OMST-Pearson, respectively.

**Figure 4:**
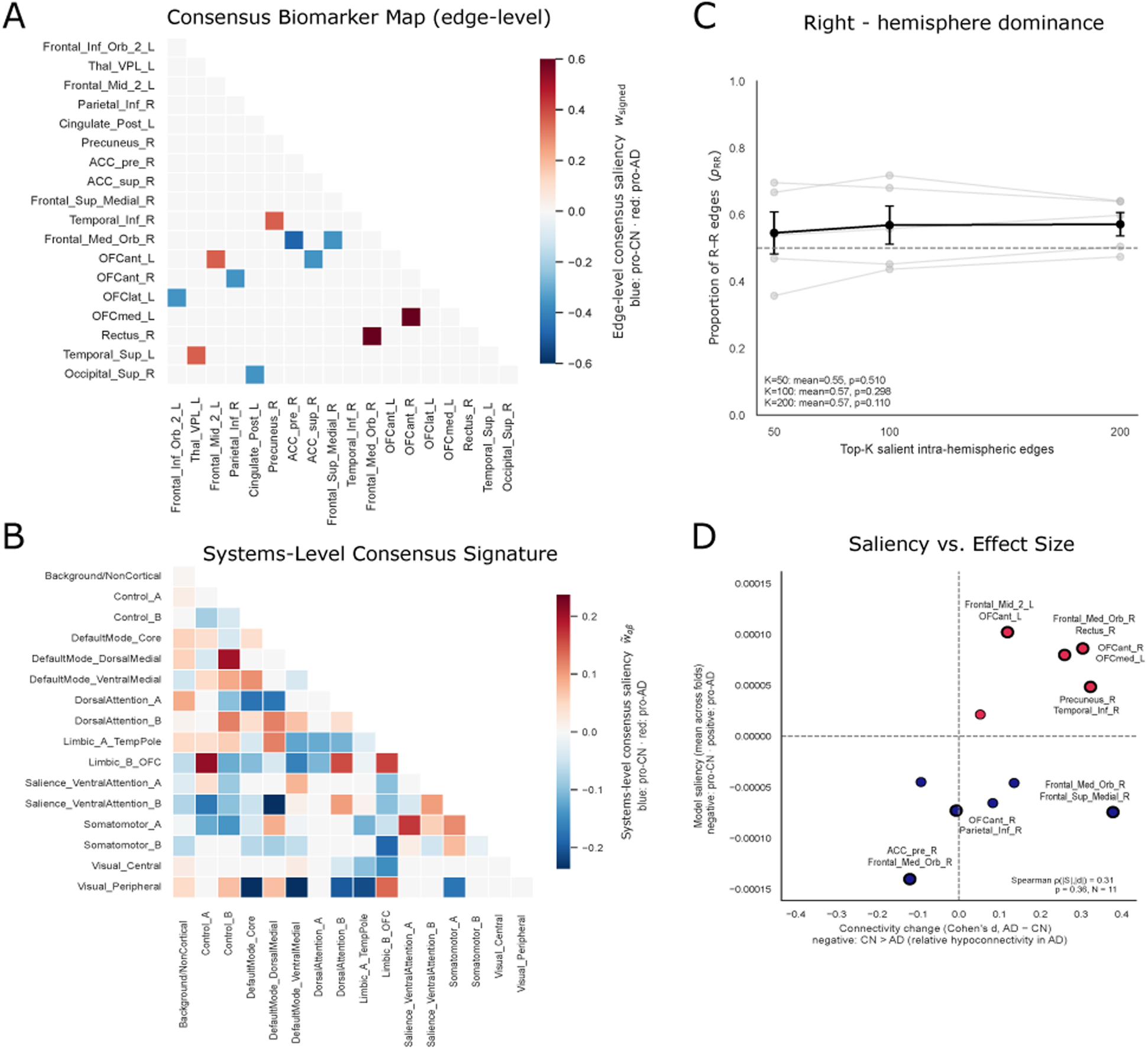
A stable AD connectivity signature with a right-lateralisation trend. **(A)** Consensus map retaining edges with high replication frequency and sign consistency, blue: pro-CN, red: pro-AD. **(B)** Systems-level signature aggregated from the consensus edges. **(C)** Lateralisation: proportion of right–right (R–R) edges among Top-K intra-hemispheric connections (bars = mean across folds; whiskers = ±1 SEM). Summary statistics and p-values from two-sided one-sample t-tests against p_RR_ = 0.5 are reported in Table 3. **(D)** Relationship between model saliency and held-out effect size within the 11-edge saliency consensus set. Here ΔS*_ij_* is the signed differential saliency (AD–CN difference of IG-based edge attributions, Eq. 5) and d*_ij_* is the held-out Cohen’s d.

**Table 3:** Hemispheric laterality of Top-. K **salient edges.** For each K, we report the mean proportion of right–right (R–R) connections among intra-hemispheric edges across outer folds, and the p-value of a two-sided one-sample t-test against hemispheric symmetry (H_0_ : p_RR_ = 0.5, n = 5 folds).

| Top-$K$ | Mean $p_{\text{RR}}$ | SD | $p$-value |
| --- | --- | --- | --- |
| 50 | 0.545 | 0.140 | 0.510 |
| 100 | 0.569 | 0.128 | 0.298 |
| 200 | 0.571 | 0.078 | 0.110 |

#### Stability and consensus across folds

For each outer fold, edges were ranked by absolute differential saliency |Δ*S_ij_*|, and the Top-*K* lists (*K* ∈{50, 100, 200}) were used to compute replication frequency (*π_ij_*) and sign consistency across folds (|*s̄_ij_*|). Despite the mean Jaccard coefficient between Top-200 sets across folds being only 0.048, using thresholds (*π_ij_* ≥ 0.6, |*s̄_ij_*| ≥ 0.6), we identified an 11-edge consensus set (Fig. 4A)(full list in Table S5). The consensus set was projected into the functional network template to obtain a consensus signature at the system level (Fig. 4B), highlighting consistent disruption involving interactions among the Default Mode, Limbic, and Visual networks.

#### Hemispheric patterns

As summarised in Table 3, the proportion of R–R connections among the most salient intra-hemispheric links was slightly above symmetry. This rightward bias was modest and did not reach conventional significance (two-sided one-sample *t*-tests against *p*_RR_ = 0.5: *K* = 50, *p* = 0.510; *K* = 100, *p* = 0.298; *K* = 200, *p* = 0.110), but it was consistent across thresholds (Fig. 4C).

#### Saliency–effect size linkage for validation

To evaluate whether the classifier merely rediscovered the largest univariate AD–CN differences, we compared model saliency Δ*S_ij_* with held-out Cohen’s *d_ij_* computed only on outer-test subjects. Within the 11-edge consensus set, |*S*| and |*d*| were only weakly related (Spearman *ρ* = 0.31, *p* = 0.36, *N* = 11)(Fig. 4D), and the consensus sets derived independently from saliency and from |*d*| showed no overlap.

When expanding the analysis to the 250 most salient non-consensus edges (*N* = 483), |*S*| and |*d*| were strongly anticorrelated (Spearman *ρ* = −0.74, *p <* 10^−80^), showing that the model assigns high saliency to edges that are not necessarily those with the largest mass-univariate differences.

A quadrant analysis in the (*d_ij_,* Δ*S_ij_*) plane (Fig. 4D) shows that: (i) edges with (*d >* 0, Δ*S >* 0) act as internal positive controls (hyperconnectivity in AD and pro-AD saliency) where the latent-space model and mass-univariate statistics agree, (ii) edges with (*d <* 0, Δ*S <* 0) show hypoconnectivity in AD but are weighted as pro–CN evidence, and (iii) edges with (*d >* 0, Δ*S <* 0) display strong hyperconnectivity in AD but reduced multivariate relevance after VAE compression. No consensus edges fell in the quadrant (*d <* 0, Δ*S >* 0).

Overall, this pattern confirms that the classifier does not simply mirror classical group differences but extracts a distributed, task-specific pattern.

#### Spatial layout of the consensus signature

The projection of the 11 consensus edges onto an MNI glass brain (Fig. 5A) shows a compact subgraph involving orbitofrontal, temporo-limbic, posterior cingulate, and occipital regions. Signed consensus weights (*w*_signed_) differentiate *pro-AD* (red) and *pro-CN* (blue) edges, with visual prominence proportional to |*w*_signed_|. The maximum absolute weight was 0.60. The consensus subgraph comprises 5 pro-AD and 6 pro-CN connections. See in Supplementary Figure S3 for an alternative visualization of network-level interactions using a chord diagram.

**Figure 5:**
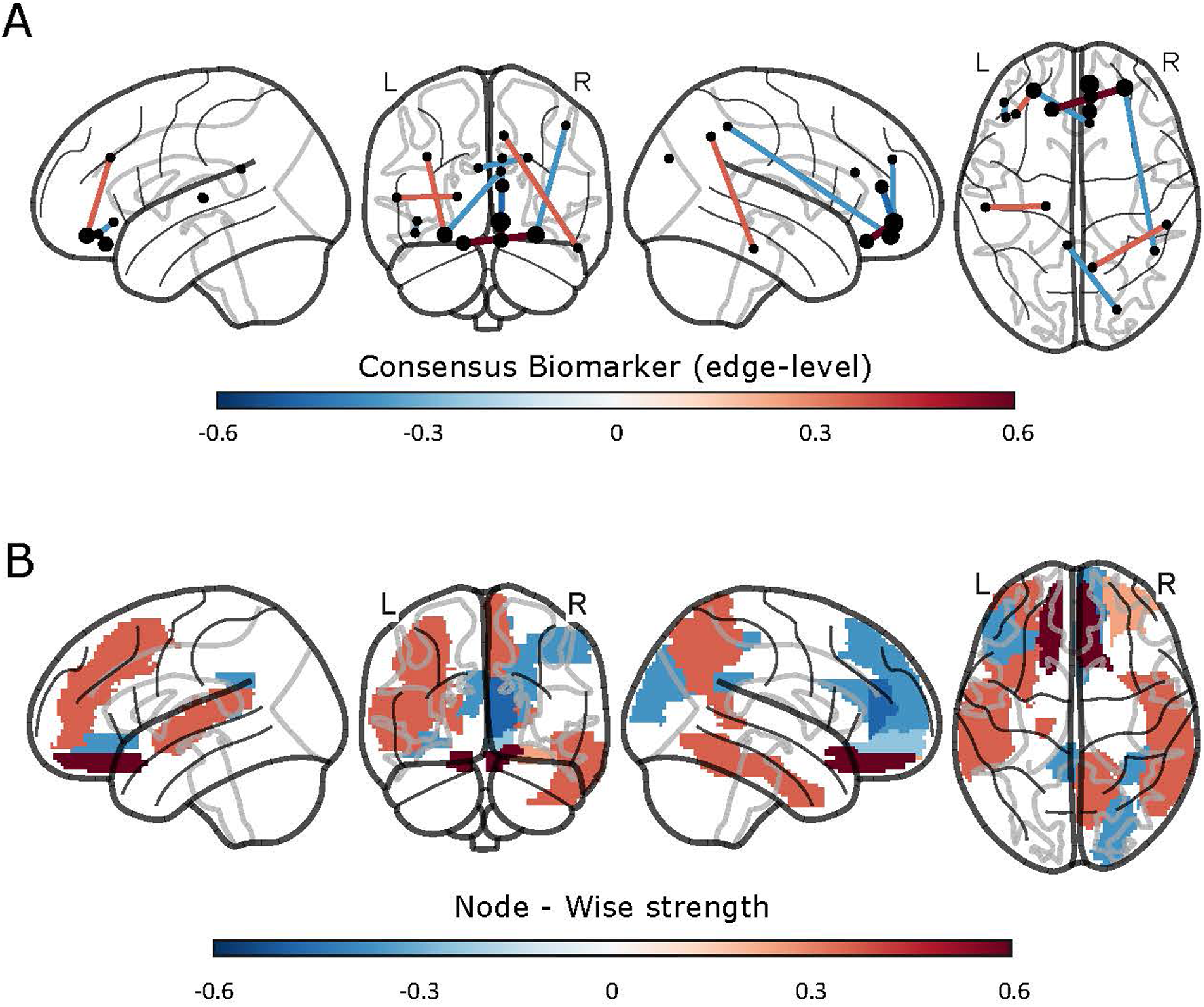
Spatial layout and nodal strength of the consensus signature. **(A)** Consensus connectome projected on a glass brain, where red (pro-AD) edges have positive AD–CN differential saliency (they increase the AD-oriented score), and blue (pro-CN) edges have negative differential saliency.**(B)** Node-wise signed strength. Blue: pro-CN, red pro-AD.

Node-wise aggregation of signed consensus weights (Fig. 5B) highlights orbitofrontal and anterior temporal ROIs as hubs with positive directional strength (pro-AD), while nodes with negative strength appear as candidate pro-CN hubs. These patterns provide a compact spatial summary of the multivariate AD–CN signature extracted by the model. Together, the edge-wise consensus map (Fig. 5A) and the nodal signed-strength pattern (Fig. 5B) provide a compact yet spatially interpretable summary of the AD-related connectivity signature learned by the model.

Overall, the integration of SHAP and IG reveals a stable, spatially structured, and multivariate connectivity signature distinct from mass-univariate AD–CN differences, and consistent with distributed fronto-limbic and posterior cortical involvement.

## 4 Discussion

In this work, we present a multichannel, explainable representation-learning framework to characterize functional connectivity alterations in Alzheimer’s disease, with a design that is readily extensible to other neurodegenerative conditions. By combining multiple complementary connectivity measures, a diagnosis-agnostic convolutional *β*-Variational Autoencoder, and a two-stage interpretability pipeline, we aimed to balance predictive performance, biological interpretability, and methodological robustness under strict cross-validation.

Our results highlight three main findings. First, a lightweight but systematic ablation analysis showed that a small subset of connectivity measures (specifically Full Pearson, OMST-Pearson, and Mutual Information *k*NN) captures most of the discriminative signal for CN–AD classification, supporting the use of compact multichannel representations over increasingly complex or dynamic features. Second, latent embeddings learned in a diagnosis-agnostic manner yielded competitive and well-calibrated classification performance when coupled with standard supervised models, despite exhibiting minimal intrinsic class separability, underscoring the value of separating representation learning from diagnostic discrimination. Third, the combination of latent-space SHAP analysis with Integrated Gradients back-projection recovered a compact and fold-stable connectivity signature, predominantly involving default-mode, limbic, and visual systems, providing a reproducible systems-level interpretation of disease-related functional alterations.

Together, these findings suggest that explainable, two-stage representation-learning approaches can offer a principled compromise between performance, interpretability, and robustness in rs-fMRI–based studies of Alzheimer’s disease, while remaining flexible to other clinical contrasts and datasets.

### 4.1 Connectivity features and multichannel representation learning

A central design choice in this work concerns the selection and combination of functional connectivity (FC) measures used to construct the multichannel connectome representation. Although a wide range of static, dynamic, and effective connectivity metrics have been proposed, their utility for disease classification depends strongly on data quality, acquisition parameters, and sample size Preti et al. [2017]. To address this empirically, we performed a systematic ablation study across seven candidate channels, spanning conventional static measures, graph-filtered representations, dynamic FC summaries, and Granger-based effective connectivity.

Across both the lightweight (FAST) and high-capacity (FULL) evaluation settings, simple static connectivity measures consistently provided the strongest diagnostic signal. In particular, full Pearson correlation and its OMST-reweighted variant emerged as the most informative individual channels, while Mutual Information contributed complementary nonlinear structure when combined with these linear metrics. In contrast, dynamic FC summaries and Granger causality measures did not improve classification performance in this dataset, either alone or when added to the best-performing static channels.

Several factors likely contribute to this result. First, the rs-fMRI data were acquired with a TR of 3 seconds, limiting sensitivity to fast temporal fluctuations in functional coupling. Under such conditions, sliding-window dynamic FC estimates are known to exhibit high variance and reduced reliability, while Granger-based measures require longer, higher signal-to-noise time series to be stable Seth et al. [2015]. In moderately sized, multi-site clinical datasets, these limitations can outweigh the theoretical advantages of richer temporal or directional models. Second, the predominance of static correlation-based measures is consistent with prior evidence that Alzheimer’s disease primarily manifests as sustained disruptions of large-scale functional coupling rather than transient connectivity states Greicius et al. [2004]; Sanz-Arigita et al. [2010]; Dennis and Thompson [2014]. In this context, OMST reweighting may further enhance disease-relevant structure by suppressing weak or redundant edges and emphasizing the backbone of the functional network.

Importantly, these findings do not preclude a role for dynamic or effective connectivity in other settings. Shorter-TR acquisitions, higher temporal signal-to-noise ratios, or longitudinal and task-based designs may provide more favorable conditions under which such features contribute meaningfully beyond static measures Lurie et al. [2020]. Future work may therefore revisit their integration within the same multichannel representation learning framework.

### 4.2 Performance, calibration, and bias control

Our two-stage pipeline yields robust CN vs. AD discrimination under fully nested cross-validation and strict leakage control. Using fold-specific latent means (256-dimensional *µ* vectors) concatenated with age and sex, Logistic Regression achieved a fold-wise mean ROC–AUC of 0.84 and mean PR–AUC of 0.86, with pooled ROC–AUC of 0.83 and a low Brier score of 0.197 (Fig. 3, Table 2). The sharp precision–recall transition and low Brier score indicate that probabilistic outputs are both discriminative and well calibrated, a property that is essential if such models are to support clinical decision-making rather than act as opaque labels. To facilitate clinical interpretation, we additionally report several standard post hoc operating points (e.g., Youden-optimal, high-sensitivity screening, and cost-sensitive thresholds) computed on pooled out-of-fold predictions (Supplementary Table S4). A central motivation for our nested design is to obtain realistic and generalizable performance estimates. Very high accuracies reported in rs-fMRI AD classification are often linked to non-nested validation or subtle forms of leakage between training and test data, while rigorous nested CV and fold-wise model selection tend to yield more conservative—and more reliable—estimates Cawley and Talbot [2010]; Varoquaux [2018]; Dadi et al. [2019]; Dinsdale et al. [2021]; Abrol et al. [2021]. In this context, ROC–AUC values around 0.8–0.85, combined with explicit calibration and interpretability, may be more compatible with eventual deployment than apparently “perfect” but potentially optimistic benchmarks.

We systematically examined potential biases in both input and latent spaces. Scanner-manufacturer decodability from the latent embeddings was moderate (k-NN balanced accuracy ≈ 0.42; chance = 1*/*3) and comparable to that obtained directly from the input connectomes. Multinomial logistic decoders reached balanced accuracies in the 0.68–0.74 range for both inputs and latents (Supplementary Tables S1–S2), indicating that the *β*-VAE preserves–and slightly concentrates–the acquisition-related structure that is already present in the raw features, rather than creating new confounding axes. Age and sex were only weakly decodable from the latent means, and none of our confound indices (latent separability, manufacturer/site decodability, demographic summaries) showed a consistent monotonic association with AD performance across folds (all Spearman correlations with outer-fold ROC–AUC had *p* ≥ 0.10; Supplementary Material S5). Together, these results suggest that acquisition and demographic factors remain detectable but do not appear to be the primary drivers of CN–AD discrimination in our framework.

### 4.3 Neurobiological interpretation

Our pipeline is explicitly designed to move from a high-capacity deep model to a small, reproducible set of connectivity features. The SHAP→Integrated-Gradients (IG) attribution path, applied to the frozen encoder and trained classifier, recovers a multivariate, fold-stable signature that is not reducible to simple mass-univariate effects.

When we enforced fold-wise stability—requiring edges to appear frequently across folds and to maintain a consistent sign, only 11 connections met the strict consensus criteria (Fig. 4B, supplementary Table S5). This 11-edge subgraph represents a highly conservative core, obtained after discarding the large majority of fold-specific top-ranked edges. The low pairwise overlap among these fold-specific edge sets (≈ 0.05) highlights the inherent instability of high-dimensional feature rankings and motivates our consensus-based approach.

From a neurobiological perspective, the consensus signature reveals a mixed pattern of relative hyper- and hypoconnectivity in AD, distributed across orbitofrontal, cingulate, temporal, parietal, and posterior midline regions (Fig. 4). This pattern is consistent with the view that Alzheimer’s disease involves both loss of functional integration in vulnerable systems and regionally specific increases in coupling that may reflect compensatory or maladaptive network reorganisation.

At the regional level, one of the most salient pro-AD edges links the right inferior temporal cortex and the right precuneus. Both regions are central to Alzheimer’s disease pathophysiology: the precuneus is a core hub of the default mode network (DMN) and among the earliest sites of amyloid and metabolic disruption, while inferior temporal regions are closely associated with memory and semantic processing deficits. Increased coupling between these regions has been reported in early or prodromal stages of AD and is often interpreted as reflecting compensatory recruitment within the posterior DMN–temporal axis.

We also observe increased connectivity involving orbitofrontal regions, particularly between medial and anterior orbitofrontal cortices and adjacent limbic structures, including the gyrus rectus. Prior studies have reported orbitofrontal hyperconnectivity in early AD and MCI, potentially reflecting compensatory engagement or loss of network segregation within limbic–prefrontal circuits Chauveau et al. [2025]; Qi et al. [2019]. While some reports describe reduced orbitofrontal connectivity in later disease stages Feng et al. [2019], the directionality observed here may be consistent with our cohort composition, which predominantly reflects baseline AD subjects from ADNI and may therefore be biased toward earlier clinical stages.

On the other hand, our model also captured a decrease in connectivity between the right anterior cingulate cortex (ACC) and medial frontal areas. Disruption of ACC-centered networks has been consistently reported in both AD and MCI and is thought to contribute to impairments in cognitive control and attentional regulation Wang et al. [2024]. At the systems level, these effects align with reduced connectivity within dorsal components of the DMN and salience-related networks, a hallmark of AD-related network breakdown Zhou et al. [2010]; Wu et al. [2011].

Beyond limbic and default-mode systems, the consensus signature highlights disconnection involving the Peripheral Visual network, the core and ventral subdivisions of the DMN, and the Limbic A temporal pole subsystem. Loss of long-range integration between visual, default-mode, and limbic networks has been previously described in AD and linked to impairments in visuospatial processing and higher-order perceptual integration Damoiseaux et al. [2012]; Krajčovičová et al. [2014].

Finally, at the network-interaction level, we observe increased connectivity between frontoparietal control networks and both dorsal-medial DMN and orbitofrontal–limbic regions. Such patterns have been interpreted as reflecting compensatory engagement of control networks in response to declining efficiency of default-mode and limbic systems Zhao et al. [2019]; Zhukovsky et al. [2022]. Importantly, while we observe a mild rightward bias in intra-hemispheric edges, this effect does not reach conventional significance and should be interpreted cautiously, consistent with a largely bilateral reorganisation process rather than strong hemispheric lateralisation.

### 4.4 Methodological implications for interpretable deep learning

Methodologically, this work illustrates how a generative, diagnosis-agnostic representation learning stage can be decoupled from diagnostic discrimination to improve robustness and interpretability in deep learning models for neuroimaging. By training the *β*-VAE independently of class labels, the model learns a smooth, data-driven functional connectivity manifold that captures inter-subject variability without being forced to align with potentially noisy diagnostic boundaries. Diagnostic discrimination is then performed in the resulting low-dimensional latent space, allowing the supervised component to remain deliberately simple.

Operating in this compact *µ*-space enables the use of linear or shallow classifiers (Logistic Regression or small MLPs), which reduces overfitting risk and substantially simplifies interpretation. In contrast to end-to-end deep classifiers, where decision logic is distributed across many layers and difficult to disentangle, this design localizes class-relevant information to a small number of latent axes that can be explicitly interrogated. This separation is a key enabler of the SHAP→Integrated Gradients attribution strategy used here.

Our framework also directly addresses several methodological concerns raised in recent reviews of deep learning for Alzheimer’s disease neuroimaging Chamakuri and Janapana [2025]. First, we adopt strictly nested cross-validation with fold-wise model selection, yielding conservative but reliable performance estimates and avoiding optimistic bias Cawley and Talbot [2010]; Varoquaux [2018]. Second, rather than training deep CNNs directly on high-dimensional fMRI time series or volumes Li et al. [2020]; Zhang et al. [2021], we use spatially structured connectivity matrices as an intermediate representation that is naturally aligned with network neuroscience and supports biologically meaningful attribution at the level of edges and systems. Third, we explicitly build an XAI layer into the pipeline by combining SHAP in latent space with Integrated Gradients back-projected to the connectome. In addition, by enforcing fold-wise stability, we move away from “black-box” predictions towards reproducible and biologically interpretable saliency maps Arya et al. [2023]; Balne and Elumalai [2021]; Lee et al. [2024]. This emphasis on stability is particularly important in high-dimensional neuroimaging settings, where attribution patterns are otherwise known to be highly variable across resampling schemes.

Taken together, our results support a shift from increasingly deeper and less transparent architectures towards rigorously validated, connectivity–aware, and explainable models. Even if this implies more modest headline accuracies, such models may offer a more credible route and scientifically grounded path toward clinically useful AI in Alzheimer’s disease.

### 4.5 Limitations and future directions

Several limitations should be considered when interpreting these results. First, although the CN/AD classification cohort (*N* = 184) is comparable to many rs-fMRI studies in Alzheimer’s disease, it remains moderate in size. While we mitigated overfitting through strictly nested cross-validation and stability analyses, external validation on independent cohorts (e.g., AIBL, OASIS) will be necessary to fully assess generalisability across scanners, protocols, and populations. Relatedly, our analyses relied on a single anatomical parcellation (AAL3), although this choice facilitates interpretability, future work should test the robustness of the consensus signature across alternative atlases and spatial resolutions.

Second, the present study is cross-sectional. As such, the identified latent axes and connectivity signatures reflect group-level differences between CN and AD at baseline rather than disease progression per se. Longitudinal extensions of the same framework could leverage the diagnosis-agnostic latent space to model trajectories of cognitive decline, MCI-to-AD conversion, effectively turning the latent space into a staging or prognosis manifold.

Third, while our explainability pipeline combines SHAP and Integrated Gradients with explicit fold-wise stability constraints, interpretability in deep learning remains an open methodological challenge. Alternative attribution strategies (e.g., layer-wise relevance propagation or perturbation-based methods) and complementary robustness tests may provide additional insights into the stability and specificity of the identified biomarkers.

Finally, acquisition-related factors such as scanner manufacturer remain partially encoded in the latent space. Although we found no evidence that these factors systematically drive CN–AD discrimination, and their influence was comparable to that present in the input connectomes, future work could incorporate explicit harmonisation or domain-adaptation strategies in latent space, particularly in large multi-site or multi-cohort studies.

Finally, acquisition-related factors such as scanner manufacturer remain partially encoded in the latent space. However, supervised performance remained robust, and we found no evidence that these confounds systematically drive AD discrimination. Future work could incorporate explicit harmonisation or domain-adaptation strategies in latent space, particularly in large multi-site or multi-cohort studies.

## 5 Conclusion

In this study, we present an explainable deep-learning framework for the analysis of resting-state functional connectivity in Alzheimer’s disease. By combining diagnosis-agnostic latent representation learning with a transparent, stability-aware attribution pipeline, our approach yields competitive and well-calibrated CN–AD discrimination while identifying a compact and reproducible connectivity signature involving default-mode, limbic, and visual systems. Rather than relying on highly complex classifiers, the framework decouples representation learning from diagnosis and supports interpretation at both the edge and network levels, revealing multivariate disease-related patterns. These results demonstrate a methodological route toward interpretable and reproducible connectivity-based biomarkers, and motivate external validation as a critical next step toward clinically meaningful tools for neurodegenerative disease research.

## Data Availability

All data produced in the present work are contained in the manuscript

https://github.com/santiblas/betavae-xai-ad

## Acknowledgments

Data used in the preparation of this article were obtained from the Alzheimer’s Disease Neuroimaging Initiative (ADNI) database (adni.loni.usc.edu). As such, the investigators within the ADNI consortium contributed to the design and implementation of ADNI and/or provided data but did not participate in the analysis or writing of this article. A complete listing of ADNI investigators can be found at: http://adni.loni.usc.edu/wp-content/uploads/how_to_apply/ADNI_Acknowledgement_List.pdf.

## Data and Code Availability

All code for preprocessing, feature extraction, modelling, and attribution, together with the AAL3→Yeo-17 mapping CSV and fold artefacts, is available at: github.com/santiblas/betavae-xaiad (commit 823b285a). A frozen release will be archived on Zenodo with a DOI; the link will be added upon publication.

## Ethics

This study includes analyses of ADNI database, a publicly available neuroimaging dataset, which was obtained in accordance with their respective institutional ethical guidelines and participant consent procedures.

## Author Contributions

MAB, DMM, and SVBL designed the study. MAB and SVBL collected and processed the data. SVBL developed the analysis code and performed the analyses. HA, DMM, and MAB supervised the work. SVBL drafted the initial manuscript, and all authors (SVBL, HA, DMM, and MAB) contributed to manuscript revision and approved the final version.

