## supplementary material for "Stable Network-Level Functional Connectivity Alterations in Alzheimer’s Disease Identified via Interpretable Latent Modelling"

#### S1 Connectivity measures: definitions and implementation details

Let $\mathbf{S} \in \mathbb{R}^{T \times Roi}$ be the subject-level matrix of ROI time series ($T$=140 time points, $Roi$=131 ROIs), with column $s_i \in \mathbb{R}^T$ the $z$-scored BOLD of ROI $i$. Each connectivity channel yields a symmetric matrix $\mathbf{C}^{(k)} \in \mathbb{R}^{Roi \times Roi}$ with zero diagonal. Unless stated otherwise, FC estimates were computed on full-length time series. All channels were z-scored on off-diagonal entries using training subjects only, ensuring commensurability across channels.

**Full Pearson correlation (Fisher-$z$ transformed).** Full Pearson correlation was computed between ROI time series and then Fisher-z transformed to stabilize variance ($\mathbf{C}^{(\text{Pearson})}$).

**OMST-Pearson** To obtain a sparse topology-preserving backbone, we applied the Orthogonal Minimum Spanning Tree with the Global Cost Efficiency (GCE) criterion (Dimitriadis et al., 2017) to the dense Fisher-$z$ matrix. The resulting binary mask was reweighted with the original Fisher-z signs to obtain a sparse signed connectivity matrix $\mathbf{C}^{(\text{OMST})}$.

**Distance correlation.** Distance correlation captures any (linear or non-linear) statistical dependence between two univariate series (Székely et al., 2007). We computed $\mathbf{C}^{(\text{dCor})}$ using the `dcor` implementation (biased, consistent estimator).

**Mutual information $k$-NN.** Mutual information between ROI time series was estimated using the Kraskov–Stögbauer–Grassberger k-nearest neighborS estimator (k=5), through the `scikit-learn`'s `mutual_info_regression` implementation for continuous variables. To reduce small-sample asymmetries of the estimator, MI was computed in both directions and averaged, yielding a symmetric nonnegative connectivity matrix $\mathbf{C}^{(\mathrm{MI})}$.

**Dynamic Functional Connectivity (dFC): mean absolute fluctuation.** To capture temporal variability in functional connectivity, we computed dynamic functional connectivity using a sliding-window approach (Allen et al., 2014). We used rectangular sliding window of length $w$=30 TR (90 s) and steps of $s$=5 TR (15 s). Let $W = \lfloor \frac{T-w}{s} \rfloor + 1$ be the number of windows, $\mathbf{R}^{(t)}$ the Pearson matrix in window $t$, and $\mathbf{A}^{(t)} = |\,\mathbf{R}^{(t)}\,|$. The mean absolute fluctuation is

$$\mathbf{C}^{(\mathrm{dFC\text{-}MAD})} = \frac{1}{W-1} \sum_{t=2}^{W} \left|\, \mathbf{A}^{(t)} - \mathbf{A}^{(t-1)} \,\right|, \qquad C_{ii}^{(\mathrm{dFC\text{-}MAD})} = 0. \tag{S1}$$

This nonnegative channel highlights edges whose absolute correlation changes rapidly over time, which has been proposed as a marker of altered network flexibility in neurodegenerative and neuropsychiatric conditions.

**Dynamic Functional Connectivity (dFC): temporal instability.** As a complementary dynamic measure, we estimated temporal instability, defined as the variance of each edge's connectivity strength across sliding windows, using the same windows as dFC mean absolute fluctuation. This channel ($\mathbf{C}^{(\mathrm{dFC\text{-}SD})}$) is nonnegative and larger for edges with high temporal variability.

**Symmetric Granger $F$-scores.** To assess directed interactions between brain regions, we computed pairwise Granger causality between ROI time series with Lag=1 (Granger, 1969). Bivariate autoregressive models were fitted to the preprocessed BOLD signals, and the resulting Granger causality estimates were symmetrized to obtain an undirected representation compatible with the remaining connectivity measures. Although interpretation of Granger causality in fMRI is limited by temporal resolution and hemodynamic confounds, it was included as a candidate metric to test whether directed information flow provides complementary diagnostic signal prior to ablation.

Together, these static, dynamic, and directed connectivity metrics were included as candidate channels to capture complementary aspects of functional brain organization prior to the ablation-based channel selection described in the main text.

#### Post-processing common to all connectivity channels

All connectivity matrices underwent identical post-processing steps prior to downstream analyses. (i) Matrices are symmetrized and diagonal elements were set to zero. (ii)

For each subject and channel, off-diagonal z-scoring was applied to ensure comparability across channels. Means and standard deviations were computed using training subjects only within each outer cross-validation fold, thereby avoiding information leakage. (iii) ROI ordering followed a consistent AAL3-to-Yeo-17 network reindexing, enabling network-level aggregation and interpretation.

Table S1: Overview of the functional connectivity measures considered, including their signedness, temporal nature (static/dynamic), and key parameters.

| Channel | Signed? | Static / Dynamic | Key params |
|---|---|---|---|
| Full Pearson (Fisher-$z$) | Yes | Static | – |
| OMST-Pearson | Yes | Static | OMST-GCE |
| Distance correlation | No | Static | – |
| Mutual information $k$NN | No | Static | $k$=5 |
| dFC: mean absolute fluctuation | No | Dynamic | $w$=30, $s$=5 |
| dFC: temporal instability | No | Dynamic | $w$=30, $s$=5 |
| Symmetric Granger $F$-scores | No | Static | $L$=1 |

### S2 $\beta$-VAE Architecture and Training

**Objective.** We learn a compact, disease-agnostic manifold of multichannel functional connectomes with a convolutional $\beta$-VAE. The loss is the ELBO with an explicit $\beta$ weight on the KL term:

$$\mathcal{L}_{\beta\text{-VAE}} = \mathbb{E}_{q_\phi(z|x)}[\log p_\theta(x|z)] - \beta D_{\mathrm{KL}}(q_\phi(z|x) \,\|\, \mathcal{N}(0, I)). \tag{S2}$$

**Input representation and normalization.** The VAE is trained with the seven channels per subject for the ablation study, and then with the three resultant channels (Full Pearson, OMST-Pearson, and Mutual Information $k$NN) for the final high-capacity VAE framework. The channels were stacked as a $N_C \times 131 \times 131$ tensor. Input normalisation and ROI reordering followed the procedures described in the main Methods and were applied fold-wise using training data only.

**Encoder and latent space.** The encoder consisted of four strided convolutional layers/blocks with GELU activations, group normalization, and dropout ($p = 0.2$), followed by a fully connected projection:

$$\text{Conv2d}(k{=}7, s{=}2) \rightarrow \text{GELU} \rightarrow \text{GroupNorm}(g{=}16) \rightarrow \text{Dropout}(p{=}0.2),$$

$$\text{Conv2d}(k{=}5, s{=}2) \rightarrow \text{GELU} \rightarrow \text{GroupNorm}(g{=}16) \rightarrow \text{Dropout}(p{=}0.2),$$

$$\text{Conv2d}(k{=}5, s{=}2) \rightarrow \text{GELU} \rightarrow \text{GroupNorm}(g{=}16) \rightarrow \text{Dropout}(p{=}0.2),$$

$$\text{Conv2d}(k{=}3, s{=}2) \rightarrow \text{GELU} \rightarrow \text{GroupNorm}(g{=}16).$$

The resulting tensor ($128{\times}8{\times}8$) is flattened (8192 units) and passed through a fully connected layer of size 2048. Two linear heads produce the network outputs: the parameters of a diagonal Gaussian latent distribution with fixed dimensionality $d_z = 256$, namely the latent mean $\mu \in \mathbb{R}^{256}$, and log-variance $\log\sigma^2 \in \mathbb{R}^{256}$. During training, latent samples were obtained via the reparameterisation trick. For all downstream analyses, only the latent means $\mu$ were used.

**Decoder.** The decoder mirrored the encoder architecture using transposed convolutions to reconstruct the input connectivity matrices. A final `tanh` activation was used, consistent with the $z$-scored input channels.

**Optimisation and schedules.** We train with AdamW (lr $1{\times}10^{-4}$, weight decay $1{\times}10^{-5}$), batch size 64, up to 2400 epochs, and *early stopping* with patience 120 on the validation ELBO. We use `CosineAnnealingWarmRestarts` ($T_0$=120, $\eta_{\min}$=$5{\times}10^{-7}$) and training uses *mixed precision*. The $\beta$ schedule is *cyclical* with $n_{\text{cycles}}$=32 and $\beta_{\max} = 2.5$; in each cycle, $\beta$ increases linearly during the first 40% and remains at $\beta_{\max}$ thereafter:

$$\beta_t = \begin{cases} \beta_{\max}\dfrac{\tau}{0.4}, & \tau < 0.4, \\ \beta_{\max}, & \text{otherwise}, \end{cases} \quad \text{with} \quad \tau = \frac{t \bmod T_{\text{cycle}}}{T_{\text{cycle}}}.$$

**Data splits and leakage control.** Within each outer cross-validation fold of the supervised pipeline, the VAE was trained diagnosis-agnostically on the full training pool only (including CN, MCI, and AD subjects), explicitly excluding the corresponding test fold. An internal validation split (20%) was used for early stopping and hyperparameter monitoring. This ensured that representation learning remained strictly leakage-free with respect to downstream classification and interpretability analyses.

**Outputs for downstream analyses.** After convergence, we freeze the encoder and extract the latent means $\boldsymbol{\mu} \in \mathbb{R}^{256}$ for each subject. These latent embeddings are the only imaging features used by the supervised classifiers and the subsequent interpretability analyses.

**Reproducibility.** All experiments were implemented in Python using PyTorch and executed on NVIDIA GPUs. The code logs software versions, random seeds, and the git commit hash, and stores fold-specific artefacts to enable full regeneration of results.

#### S3 Supervised classification pipeline

This section provides implementation details for the supervised classification stage described in the main Methods. Classification was performed exclusively on the CN vs. AD subset using fold-specific latent embeddings derived from the $\beta$-VAE.

**Concrete run configuration.** Unless otherwise stated, results correspond to the following configuration:

- **Features:** VAE latent means $\boldsymbol{\mu} \in \mathbb{R}^{256}$ concatenated with Age and Sex.
- **Classifiers:** Logistic Regression, RBF-SVM, XGBoost, LightGBM, and a small MLP.
- **Cross-validation:** Nested 5×5 CV (5 outer folds; 5 inner folds for model selection).
- **Optimisation:** Optuna (TPE sampler, MedianPruner), maximising ROC–AUC.
- **Class imbalance:** Class weighting only (no synthetic oversampling such as SMOTE).
- **Calibration:** Isotonic calibration where supported.
- **Reproducibility:** Random seed = 42; artefacts saved per fold; git hash logged.

**Task and features.** For each subject, the 256-dimensional latent mean vector $\boldsymbol{\mu}$ from the corresponding fold-specific encoder was concatenated with Age and Sex to form the feature vector. Latent representations were extracted from encoders trained exclusively on the non-test pool of each outer fold (CN/MCI/AD), ensuring strict separation between representation learning and evaluation.

**Algorithms.** We evaluated five complementary classifier families spanning linear, kernel-based, ensemble, and neural-network approaches: regularised Logistic Regression (liblinear), RBF-SVM, XGBoost, LightGBM, and a shallow MLP. Implementations followed scikit-learn, with native libraries used for gradient boosting.

**Preprocessing and pipelines.** All steps were encapsulated in a single `imblearn` *Pipeline*. Non-tree models (Logistic Regression, SVM, MLP) used `StandardScaler`, while tree-based models operated on unscaled features. Class imbalance was addressed using class weighting where supported. Synthetic oversampling methods (e.g., SMOTE) were not used in any reported experiment. Probability calibration (isotonic) was applied to SVM and LightGBM via `CalibratedClassifierCV`. All preprocessing, calibration, and model fitting occurred strictly within inner training folds.

**Hyperparameter optimization.** For each classifier and outer fold, hyperparameters were optimised using `OptunaSearchCV` with a TPE sampler and MedianPruner. The inner cross-validation used 5 stratified folds and optimised ROC–AUC. Searches were capped at a maximum of 100 trials or 30 minutes per classifier. Standard parameter ranges were explored (e.g., $\{C, \gamma\}$ for SVM; learning rate, depth, and regularisation for boosting; learning rate and weight decay for MLPs). GPU acceleration was enabled for XGBoost and LightGBM when available.

**Evaluation.** Performance was assessed on each outer test fold using ROC–AUC (primary metric), PR–AUC, balanced accuracy, sensitivity, specificity, and $F_1$ score. Final results are reported as mean $\pm$ standard deviation across outer folds. Only outer-test predictions were used for performance reporting and downstream analyses.

**Leakage control.** The VAE encoder, preprocessing steps, and classifiers were all trained in a fold-specific manner excluding outer-test subjects, ensuring a fully leakage-free evaluation throughout the pipeline.

### S4 End-to-end workflow (summary)

Figure 1 summarises the end-to-end analysis pipeline. Within each outer cross-validation fold, multichannel functional connectivity matrices are computed from preprocessed rs-fMRI data and used to train a diagnosis-agnostic convolutional $\beta$-VAE on the non-test pool (including MCI subjects). The encoder is then frozen and latent mean vectors are extracted for all CN and AD subjects in that fold.

Supervised classifiers are trained on the fold-specific latent embeddings concatenated with age and sex using a nested inner cross-validation loop for hyperparameter selection. Model explanations are obtained by combining SHAP values in latent space with Integrated Gradients back-projected to the input connectivity matrices. Stability across folds is assessed via replication frequency and sign consistency, yielding a compact and reproducible connectivity signature.

Algorithm 1 summarizes step-by-step the fold-wise procedure.

**Algorithm 1** Nested cross-validation scheme for leakage-free training and attribution per outer fold

1: Split data into $F$=5 outer folds; for fold $f$: define test set $\mathcal{T}_{\text{test}}^{(f)}$ and training set $\mathcal{U}_{\text{train}}^{(f)}$
2: Build representation pool $\mathcal{P}_{\text{VAE}}^{(f)} = \mathcal{U}_{\text{train}}^{(f)} \cup$ all MCI
3: Fit per-channel $z$-**score scaler** on off-diagonals of $\mathcal{P}_{\text{VAE}}^{(f)}$; save params
4: Train $\beta$-VAE on $\mathcal{P}_{\text{VAE}}^{(f)}$ (cyclic $\beta$, AdamW); freeze encoder $E_{\phi}^{(f)}$
5: Encode $\boldsymbol{\mu}(x) \in \mathbb{R}^{256}$ for $\mathcal{U}_{\text{train}}^{(f)}$ and $\mathcal{T}_{\text{test}}^{(f)}$
6: Inner CV on $\mathcal{U}_{\text{train}}^{(f)}$ with `imblearn` pipeline
7: Evaluate best pipeline once on $\mathcal{T}_{\text{test}}^{(f)}$; store metrics
8: Compute SHAP values for each latent feature plus age and sex; and group-contrast weights $w_\ell$ (Eq. 1)
9: Integrated Gradients to obtain AD–CN *differential saliency* $\Delta S_{ij}^{(f)}$ (Eq. 4 and Eq. 5)
10: Stability and consensus: Top-$K$ edges per fold f to compute replication frequency and mean signed direction across folds

### S5 Connectivity channel ablation

To identify a compact yet informative set of connectivity channels for representation learning, we performed a greedy forward-selection ablation over the seven candidate measures: Full Pearson (Fisher-$z$), OMST–Pearson, Distance Correlation, Mutual Information $k$NN, dynamic FC: mean absolute fluctuation, dynamic FC: temporal instability (across-window standard deviation), and symmetric Granger $F$-scores.

To identify a compact yet informative set of connectivity channels for our VAE, we performed a greedy forward selection analysis over seven candidate channels: Full Pearson (Fisher-$z$), OMST (GCE) reweighted by signed Fisher-$z$, Distance Correlation (dCor), Mutual Information (KSG, symmetric), dynamic FC mean absolute fluctuation (dFC-MAD), dynamic FC across-window standard deviation (dFC-SD), and symmetric Granger $F$-scores (lag 1). This procedure was conducted under a resource-bounded **FAST configuration** designed for rapid evaluation, which differs from the FULL configuration used for the final results.

**Protocol.** A lightweight FAST configuration was used to rank channels efficiently, prioritising relative performance over absolute accuracy. This configuration employed a reduced-capacity $\beta$-VAE and a fixed Logistic Regression classifier, evaluated with 3-fold outer cross-validation using mean ROC–AUC. No hyperparameter tuning or synthetic oversampling was applied.

The selection process began by training and evaluating a separate model for each of the seven candidate channels individually. The channel yielding the highest ROC-AUC was selected as the starting point. Subsequently, in an iterative fashion, each of the remaining channels was added one by one to the current best set, and the new combination was evaluated.

**Results.** Single-channel analysis showed that Full Pearson was the strongest individual predictor (mean ROC–AUC = $0.75 \pm 0.02$). Greedy forward selection revealed that performance peaked with the two-channel combination **Full Pearson + OMST–Pearson** (mean ROC–AUC = $0.79 \pm 0.03$ SE; Fig. 2). Applying the standard one-standard-error (1-SE) rule identified this two-channel model as the most parsimonious choice under the FAST configuration.

**Transition to the FULL model.** The FAST ablation provides a conservative estimate of channel importance, prioritizing simplicity. When re-evaluated under the full-capacity pipeline used in the main analyses (deeper $\beta$-VAE, nested hyperparameter optimisation, multiple classifier families), adding **Mutual Information** to the two-channel set yielded a consistent performance improvement. Consequently, all final models were trained on the three-channel tensor comprising Full Pearson, OMST–Pearson, and Mutual Information $k$NN, to maximize the model's predictive power.

### S6 Quantitative and Visual Validation of the Latent Space

We report supplementary quantitative and visual analyses that support the main-text characterization of the learned latent space. Table S2 summarizes metrics characterizing class separability, confound (scanner manufacturer) decodability, and intrinsic dimensionality of the learned latent space. Table S3 reports a more detailed analysis of scanner-manufacturer decodability from both the input connectomes and the latent means. Figure S1 provides a complementary visual inspection of this consistency using UMAP projections

Consistent with the main results, class separability in latent space was negligible (mean silhouette $0.005 \pm 0.007$), and the effective dimensionality was substantially lower than the nominal latent size (participation ratio $\approx 20$). Scanner-manufacturer labels were decodable from latent embeddings with moderate balanced accuracy ($0.418 \pm 0.060$; chance $\approx 0.33$), comparable in magnitude to decodability from the normalized input connectomes (Table S3). These results confirm that acquisition-related structure is preserved at a measurable level in the latent space and can be explicitly quantified in downstream analyses.

Table S2: Quantitative Quality Control of the Latent Space. Metrics were computed on the standardized latent embeddings for the test set of each outer fold. Class Separability is the Silhouette Score on a 20D PCA subspace. Confounder Effect is the balanced accuracy of a k-NN classifier predicting the scanner manufacturer (chance $\approx$ 0.33). Effective Dimensions is the Participation Ratio (PR). PCA Dims (90% Var) is the number of components explaining 90% of variance.

| Fold | Class Separability (Silhouette) | Confounder (Manufacturer Acc.) | Effective Dimensions (PR) | PCA Dims (90% Var) |
|---|---|---|---|---|
| 1 | 0.011 | 0.500 | 20.973 | 23 |
| 2 | 0.003 | 0.333 | 20.678 | 23 |
| 3 | 0.013 | 0.400 | 18.711 | 22 |
| 4 | -0.005 | 0.435 | 20.370 | 22 |
| 5 | 0.001 | 0.420 | 20.432 | 22 |
| **Mean** | 0.005 | 0.418 | 20.233 | 22.4 |
| **SD** | 0.007 | 0.060 | 0.883 | 0.5 |

Table S3: Scanner-manufacturer decodability before and after VAE encoding. Balanced accuracy of a multinomial Logistic Regression decoder predicting the scanner manufacturer from either the z-scored input connectomes or the VAE latent means $\mu$ (5 outer folds). Chance is $1/3 \approx 0.33$ (three manufacturers); $\Delta\text{BA} = \text{BA} - 1/3$. Values are reported as mean $\pm$ s.d. across folds.

| Representation | BA (mean $\pm$ s.d.) | Chance | $\Delta$BA (mean $\pm$ s.d.) |
|---|---|---|---|
| Connectome (norm) | 0.678 $\pm$ 0.058 | 0.333 | 0.344 $\pm$ 0.058 |
| Latent $\mu$ | 0.735 $\pm$ 0.054 | 0.333 | 0.402 $\pm$ 0.054 |

### S7 Post hoc classification operating points

For completeness, we report in Table S4 several standard *post hoc* decision thresholds computed on the pooled ($N = 184$) out-of-fold predictions. These operating points were not used for model selection or tuning.

Table S4: Post hoc operating points computed on pooled (N=184) out-of-fold predictions. Values are derived from the final Logistic Regression model.

| **Strategy** | **Description** | **Threshold** | **Se (AD)** | **Sp (CN)** | **PPV** | **NPV** |
|---|---|---|---|---|---|---|
| Youden Index | Max (Se + Sp - 1) | 0.500 | 0.800 | 0.775 | 0.792 | 0.784 |
| Screening (Se$\geq$ 0.85) | Max Sp s.t. Se$\geq$ 0.85 | 0.472 | 0.853 | 0.640 | 0.717 | 0.803 |
| Cost-based ($C_{fn}$=3) | Min Cost | 0.456 | 0.874 | 0.607 | 0.703 | 0.818 |

*Se*: Sensitivity. *Sp*: Specificity. *PPV/NPV*: Predictive Values. Counts for Youden: $TP = 76, FN = 19, TN = 69, FP = 20$.

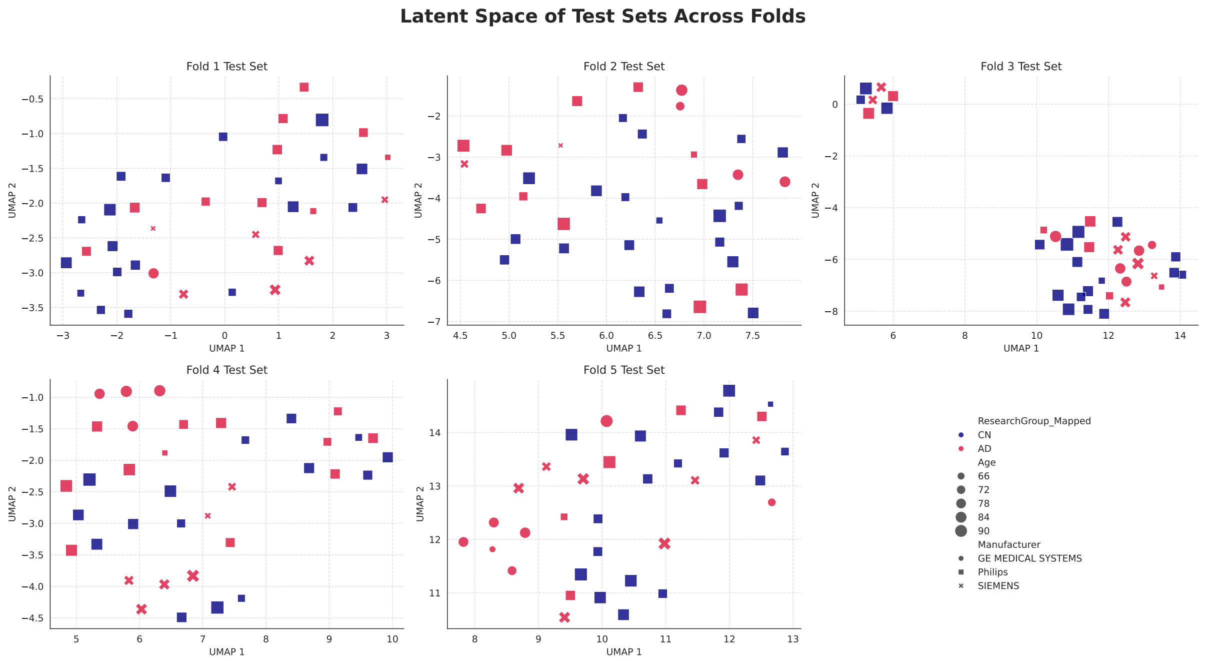


Figure S1: Latent space visualization of test sets across folds. Each panel shows the UMAP projection of latent embeddings from the held-out test subjects of the corresponding cross-validation fold, ensuring a leakage-free assessment of generalisation. Each point represents a subject, where color denotes the diagnostic group (AD/CN), marker shape indicates the scanner manufacturer (a potential confound), and marker size is proportional to the subject's age. A consistent coarse structure is observed across folds. As expected from the low silhouette values, diagnostic groups are not cleanly separated in two dimensions, and no trivial segregation by scanner manufacturer is observed.

### S8 Global latent-space explanations (SHAP)

To support the interpretability pipeline described in the main Methods, we computed global SHAP explanations for the Logistic Regression (LOGREG) head operating on the VAE latent space. LOGREG was selected as the reference classifier because it achieved the highest mean ROC–AUC across outer folds (Table 2) and provides a transparent linear decision function.

For each outer fold, SHAP values were computed on held-out test subjects using `LinearExplainer`, with a train-only background sample (100 subjects). The feature space comprised the latent means $\boldsymbol{\mu}$ concatenated with Age and Sex. To assess whether demographic covariates influenced the ranking of latent features, we compared a *frozen* regime (Age and Sex fixed to training-set statistics) with an *unfrozen* regime. Latent-only global importance profiles (mean |SHAP|) were identical across regimes in all folds (Pearson $r = 1.0$), confirming that latent rankings are robust to covariate handling.

Figure S2 shows representative SHAP summaries from one outer fold. Age consistently emerges as the most influential feature, followed by Sex and a sparse subset of latent coordinates. These results motivate the construction of the AD-oriented latent direction used for back-projection with Integrated Gradients in the main interpretability analysis.

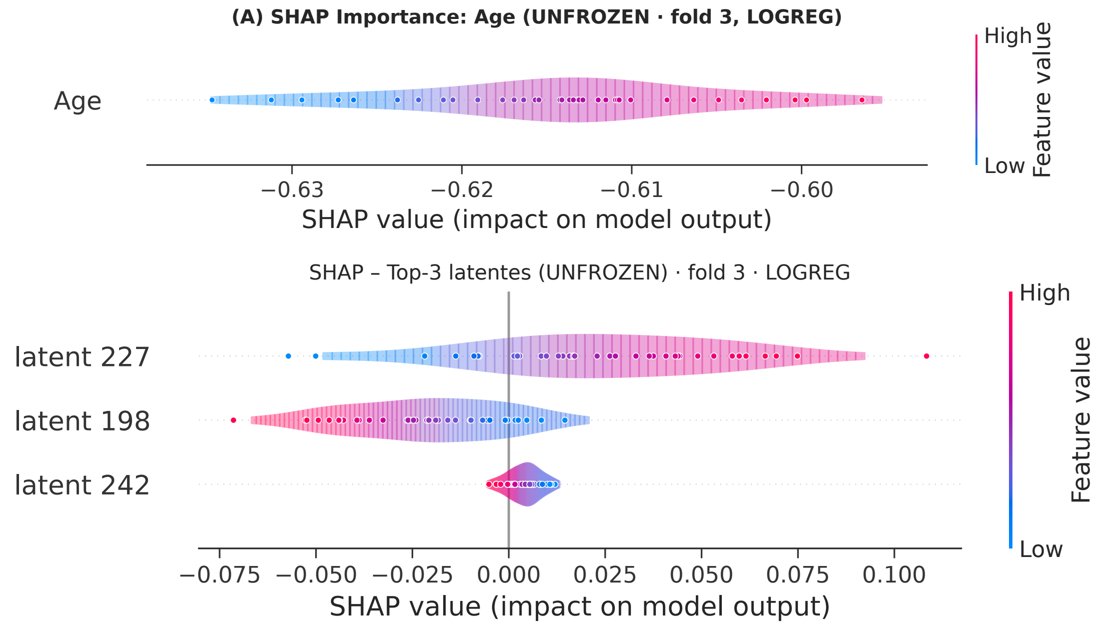


Figure S2: **Global SHAP explanations in latent space (LOGREG).** (**A**) Violin+beeswarm plot of SHAP contributions for *Age* on held-out test subjects from a representative outer fold (fold 3). (**B**) SHAP summaries for the three latent coordinates with highest global importance (latent 242, 227, and 198; ranked by mean |SHAP| across folds). Together, the panels illustrate the hierarchy *Age* > *Sex* > sparse latent coordinates in the LOGREG decision function, consistent across folds and robust to covariate handling.

### S9 Consensus Connectivity Signature

Table S5: Consensus Connectivity Signature. List of the 11 stable edges identified by the model (replication frequency $\pi_{ij} \geq 0.6$ and sign consistency $|\bar{s}_{ij}| \geq 0.6$. ROI names follow the AAL3 atlas. The network pair column reports the functional system assignment of each ROI based on the refined Yeo-17 mapping.

| **Region 1** | **Region 2** | **Network Pair (Refined)** | $\pi$ | $\lvert\bar{s}\rvert$ |
|---|---|---|---|---|
| Frontal_Med_Orb_R | Rectus_R | Limbic_B_OFC — Limbic_B_OFC | 0.60 | 1.00 |
| OFCmed_L | OFCant_R | Limbic_B_OFC — Limbic_B_OFC | 0.60 | 1.00 |
| Frontal_Med_Orb_R | ACC_pre_R | Limbic_B_OFC — DefaultMode_VentralMedial | 0.80 | 0.60 |
| Temporal_Inf_R | Precuneus_R | Limbic_A_TempPole — DefaultMode_DorsalMedial | 0.60 | 0.60 |
| OFCant_L | Frontal_Mid_2_L | Limbic_B_OFC — Control_A | 0.60 | 0.60 |
| Thal_VPL_L | Temporal_Sup_L | Background/NonCortical — Somatomotor_B | 0.60 | 0.60 |
| Occipital_Sup_R | Cingulate_Post_L | Visual_Peripheral — DefaultMode_Core | 0.60 | 0.60 |
| Frontal_Med_Orb_R | Frontal_Sup_Medial_R | Limbic_B_OFC — DefaultMode_VentralMedial | 0.60 | 0.60 |
| OFCant_R | Parietal_Inf_R | Limbic_B_OFC — Control_B | 0.60 | 0.60 |
| OFClat_L | Frontal_Inf_Orb_2_L | Limbic_B_OFC — Background/NonCortical | 0.60 | 0.60 |
| ACC_pre_R | ACC_sup_R | DefaultMode_VentralMedial — DefaultMode_VentralMedial | 0.60 | 0.60 |

### S10 Network-Level Saliency Visualization (Chord Diagram)

To complement the systems-level analysis in Fig. 5A, Fig. S3 provides an alternative visualization of network-level interactions using a chord diagram. This plot summarizes the flow of differential saliency (AD vs. CN) between functional networks (based on the refined Yeo-17 template), collapsing information across all ROI-level connections within each network pair.

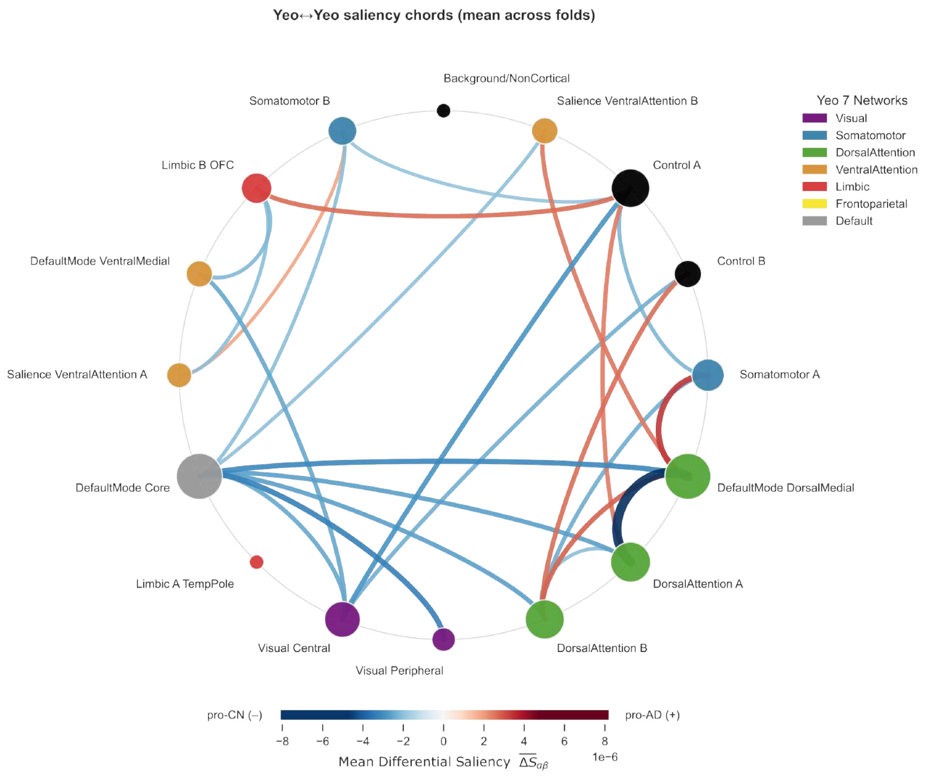


Figure S3: Chord diagram of network-level saliency (aggregate). Chords represent the aggregated differential saliency (AD–CN) between functional networks. Chord thickness is proportional to the summed mean absolute saliency ($|\Delta S|$) across all ROI-to-ROI connections linking each network pair, averaged across folds. The diagram highlights prominent intra- and inter-network interactions, with strongest contributions involving the Default Mode, Limbic, and Visual systems.
